# Naming Performance in Bilinguals with Alzheimer’s Disease and Mild Cognitive Impairment

**DOI:** 10.64898/2026.03.23.26349075

**Authors:** María Sainz-Pardo, Mireia Hernández, Anna Suades, Montserrat Juncadella, Jordi Ortiz-Gil, Lidia Ugas, Isabel Sala, Alberto Lleó, Marco Calabria

## Abstract

**Introduction:** There is consistent evidence of a disadvantage in bilinguals’ speech production compared to monolinguals in healthy individuals, but studies investigating this phenomenon in clinical populations such as Mild Cognitive Impairment (MCI) and Alzheimer’s Disease (AD) are scarce. Given that both clinical groups are characterized by word-finding difficulties, understanding how bilingualism influences speech production in these populations is essential.

**Objective:** To investigate the effect of bilingualism on dominant-language speech production in individuals with MCI and AD relative to cognitively unimpaired (CU) older adults.

**Methods:** Early and highly proficient Catalan–Spanish bilinguals (active bilinguals) were compared to Spanish-dominant speakers with low proficiency in Catalan (passive bilinguals) using a picture-naming task. The study included 58 CU older adults, 66 patients with AD, and 124 individuals with MCI. Reaction times, accuracy, and error types were collected in the naming task in each individual’s dominant language.

**Results:** First, we observed an advantage for active bilinguals compared to passive bilinguals, as indexed by faster responses, particularly for cognate words. Second, active bilinguals with MCI exhibited a disadvantage, making more naming errors than passive bilinguals with MCI, especially for non-cognates, including a higher incidence of cross-language intrusions and anomia. Third, passive bilinguals with MCI and AD showed more semantic errors than active bilinguals.

**Discussion:** Disadvantages in naming are discussed in terms of predictions from cognitive and linguistic theories, whereas potential advantages of speaking a second language are considered as a protective factor, consistent with frameworks such as cognitive reserve.

## Introduction

Word-finding deficits are a common feature of cognitive decline associated with aging and neurodegenerative disorders, such as Alzheimer’s disease (AD). While many studies have focused on understanding these deficits in monolingual speakers ^1–4^, fewer have investigated them in bilingual populations with AD (for a review, see 5). Of particular interest is the comparison between these two language groups, given the reported ‘bilingual disadvantage’ associated with speaking a second language. Although the nature of such disadvantage in healthy individuals has been explained by different hypotheses ^6–15^, how it is modulated by neurodegeneration and second language use remains little explored.

In the present study, we investigated this issue by comparing the naming performance of two groups of speakers with differing bilingual experiences: early and highly proficient Spanish-Catalan bilinguals (active bilinguals), and Spanish-dominant speakers with passive exposure to Catalan (passive bilinguals). To assess the impact of neurodegeneration, we examined the performance of cognitively unimpaired (CU) older adults, patients with Mild Cognitive Impairment (MCI), and patients with Alzheimer’s disease (AD). Both clinical populations are known to experience word-finding deficits, which are thought to primarily involving lexical ^11,13,16^ processes (access deficits) at earlier stages, and later affecting representational processes (storage deficits) (e.g., 2,3).

Beyond the theoretical motivation to understand the nature of lexical retrieval deficits in individuals with diverse language experiences, these studies are critically important for clinical practice in linguistically diverse populations. The number of people who speak more than one language is steadily increasing worldwide; at the same time, the global prevalence of neurodegenerative disorders is rising due to population aging ^17^. Consequently, examining the relationship between bilingualism and language disorders is particularly important for addressing future challenges in neuropsychology, as such disorders may manifest differently depending on a patient’s language background and use ^18^.

### The bilingual disadvantage and the impact of life-long bilingualism on speech production

Although managing two languages often appears effortless for individuals who use both in daily life, scientific research has documented a bilingual disadvantage relative to monolinguals. This disadvantage is reflected in language performance, even in the dominant language, particularly on tasks involving lexical access, such as picture naming and verbal fluency: bilinguals are slower than monolinguals and experience more tip-of-the-tongue phenomena (for reviews, see 19,20).

From a theoretical perspective, two main accounts have been proposed to explain differences in speech production performance between bilinguals and monolinguals.

According to lexical inhibition accounts, bilingual speakers must inhibit their unintended language to activate the intended one, which results in a slower access to their dominant language ^7,21,22^. Similarly, the competition account proposes that the non-target language remains activated, requiring additional effort to inhibit it and solve the conflict between cross-language competitors. In this view, the bilingual disadvantage arises from the competition between the two languages ^23^, since lexical representations in both languages are activated simultaneously, even when speakers are required to use only one of them ^24,25^.

An alternative account is the frequency-lag hypothesis ^10,26^, which relies on the idea that bilingual individuals speak each of their languages less often than monolingual speakers. Consequently, reduced frequency of language use may account for the lexical retrieval disadvantage often observed in bilinguals in their dominant language. Additionally, this hypothesis posits a parallel between overall language use frequency and word frequency. On this basis, it predicts that low-frequency words are more vulnerable in bilingual than in monolingual speakers during picture-naming tasks, leading to greater retrieval costs relative to high-frequency words ^10,27^.

Potential differences in word production and other verbal tasks between monolingual and bilingual speakers have also been examined in the context of aging, with mixed findings. Several studies reported a bilingual disadvantage in naming performance among cognitively impaired (CU) older bilinguals ^10,16,28–30^, consistent with similar disadvantages observed in younger bilinguals ^19^. In contrast, other work has reported advantages for bilinguals over monolinguals on verbal fluency tasks ^31^. Moreover, language use and dominance appear to modulate the magnitude of naming disadvantages in bilinguals relative to monolingual peers^32,33^.

Additionally, the linguistic cost associated with bilingualism appears to be sensitive to the specific linguistic demands of the task. For example, Friesen et al. ^34^ compared monolingual and bilingual speakers across the lifespan using category and letter fluency tasks. Their results revealed a stable pattern of performance in letter fluency across age groups, but an age-related decline in category fluency. Notably, older bilinguals produced more words than monolinguals in the letter fluency condition, whereas no group differences were observed in the category fluency condition.

### Word-finding deficits in bilingual and monolingual patients with MCI and AD

Although deficits in episodic memory and executive functions are considered the hallmark symptoms of AD and MCI ^35–38^, language abilities are also compromised from the early stages of the disease. In monolingual speakers, impairments may particularly affect lexical retrieval (naming and verbal fluency), but also sentence- and word-level comprehension and production at discourse level ^2,3,39^.

Within the domain of word production, lexical retrieval deficits in monolinguals have been primarily attributed to impairments of lexical access and/or access to the phonological lexicon, but also to impaired access to semantic representations rather than to a degradation of semantic knowledge itself ^40–42^. In bilinguals, naming performance has been investigated in individuals with MCI and AD, with evidence of similar degrees of decline across both languages in highly proficient bilinguals (e.g., 44,45). In contrast, less balanced bilinguals may show different patterns of language decline ^5,45^.

However, only a limited number of studies have directly compared language performance between monolinguals and bilinguals diagnosed with MCI or AD in their dominant language, and the findings remain mixed and task-dependent (e.g., fluency vs. naming). For example, Gard et al.^46^ compared cognitively unimpaired and individuals with MCI using both the Boston Naming Test and verbal fluency tasks, and found that monolinguals outperformed bilinguals on naming, but not on verbal fluency. In contrast, Anderson et al.^28^ reported evidence of a disadvantage in verbal fluency that was dependent on disease stage: although no significant differences were observed between monolingual and bilingual individuals with MCI, bilinguals with AD showed significantly poorer performance on letter fluency tasks.

In contrast, other studies have found no significant differences between monolinguals and bilinguals with MCI or AD on the same verbal tasks ^47,48^, or have reported task-dependent effects. Specifically, Bialystok et al.^47^ observed a decline in performance over time in both MCI patients (on letter fluency and category switching) and AD patients (on category fluency), regardless of language background. Conversely, Costumero et al.^49^ reported that bilingual patients with amnestic MCI showed a slower rate of decline than monolinguals on letter fluency tasks over time.

Overall, the current evidence does not allow for a clear conclusion regarding a disadvantage of bilingualism in patients with MCI and AD, highlighting the need for further research to clarify the nature of bilingualism’s effects on lexical retrieval in the dominant language.

### The present study

While previous studies in healthy individuals have examined the bilingual effect by comparing bilingual and monolingual speakers, our study takes a different approach by comparing active and passive bilinguals. To investigate whether daily use of two languages across the lifespan can modulate lexical access in individuals with MCI and AD, we focused on the naming performance of individuals who had been immersed in two distinct bilingual contexts: active versus passive language use.

These two language profiles are predominantly found in the sociolinguistic context of Catalonia, where Catalan and Spanish are co-official languages. As in our previous studies ^43,44^, we defined active bilinguals as those who acquired both languages early and simultaneously (before age 6), with high proficiency in both languages and balanced use of Catalan and Spanish. In contrast, passive bilinguals are Spanish-dominant speakers with passive exposure to Catalan and limited spoken proficiency. Comparing these two language groups is critical to determine whether active use of a second language plays a key role in preserving lexical retrieval in bilingual individuals with neurodegenerative diseases.

In particular, we compared participants’ performance on a picture naming task in their dominant language, focusing on naming latencies, accuracy, and error type. While the first two measures provide a more quantitative view of the phenomenon, the analysis of error types offers insight into potential qualitative differences in the underlying mechanisms of word-finding difficulties between the two bilingual groups. Additionally, we assessed the word frequency effect (i.e., the difference in performance between high- and low-frequency words), which provides further information about the relationship between language use and word-finding deficits.

First, based on the reported bilingual disadvantage, we hypothesized that active bilingual participants would show slower and less accurate lexical retrieval during the picture-naming task than passive bilinguals when tested in their dominant language. These expected findings would align with both the competitive account and the frequency-lag hypothesis, being consistent with the results reported by de Bruin et al.^50^ for active and passive bilinguals. Second, the difference in naming performance between active and passive bilinguals is expected to become more pronounced from MCI to AD stages, as word-finding difficulties worsen with disease progression ^42,51,52^.

Second, we examined the effect of word frequency, since it has been shown to be modulated by bilingualism. Previous studies have reported a larger word frequency effect in older bilingual adults compared to their monolingual peers ^29,53^.

Based on prior literature, we expect an increasing word frequency effect across from CU older adults to MCI and AD, which is predicted to be larger in active than in passive bilinguals, reflecting the expected bilingual disadvantage in retrieving low-frequency words. Large frequency effects are also anticipated in AD, consistent with findings showing that low-frequency words are disproportionately impaired in both monolinguals compared to healthy individuals ^54,55^ and bilinguals with AD compared to MCI ^44^.

Third, we examined the effect of cognate status, defined as the degree of overlap between the orthographic and phonological representations of a word across languages. Previous studies have shown that, beyond word frequency, cognate status is a key determinant of language performance in bilinguals with AD and MCI ^43,44^, with cognates being less affected by these conditions than non-cognates across languages. However, although a cognate advantage has also been reported in healthy individuals ^56,57^, this facilitatory effect of cognates can turn into interference in certain contexts, such as language-switching or sentence-processing tasks ^58,59^. Therefore, we might expect differential effects for cognates and non-cognates when comparing naming performance in active bilinguals naming in their dominant language.

Regarding error types, omissions and semantic errors are expected to be the most frequent, based on well-documented lexico-semantic deficits in MCI and AD ^42,60–62^. Their prevalence is predicted to increase from MCI to AD, reflecting progressive deterioration of lexico-semantic retrieval. These error types are anticipated to show a similar pattern across language groups, as they are not expected to be influenced by bilingualism. Any differences in the distribution of errors between active and passive bilinguals would likely reflect differential decline of the lexico-semantic network associated with the use of the second language.

Additionally, we predict a higher proportion of cross-language intrusions (i.e., when the target is named in the non-target language) in active bilinguals compared to passive bilinguals. This may reflect either a decline in the ability to control both languages ^63^ or a compensatory strategy in response to retrieval failure.

## Methods

### Participants

248 participants took part in the study who were recruited from different institutions in Catalonia (Spain): Hospital de Bellvitge (L’Hospitalet de Llobregat), Hospital General de Granollers (Granollers), and Hospital de la Santa Creu i Sant Pau (Barcelona). This investigation was carried out under the approval of ‘Parc de Salut MAR’ Research Ethics Committee under the reference number 2014/6003/I.

124 patients with MCI (M/F = 59/65) and 66 patients with AD (M/F = 21/45) were included in the study. Additionally, 58 CU older adults (M/F = 18/40) served as control participants (see Table 1).

**Table 1.**
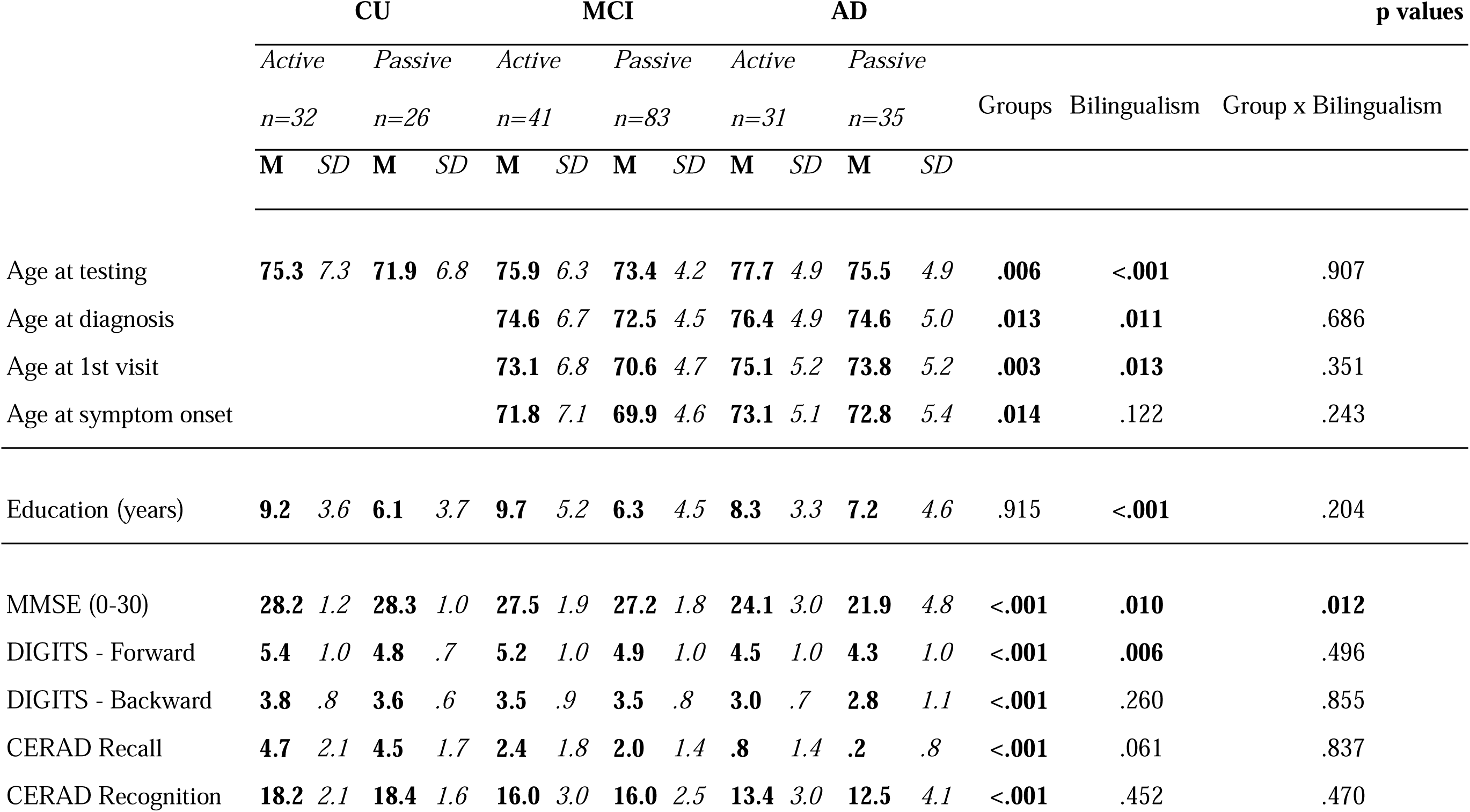
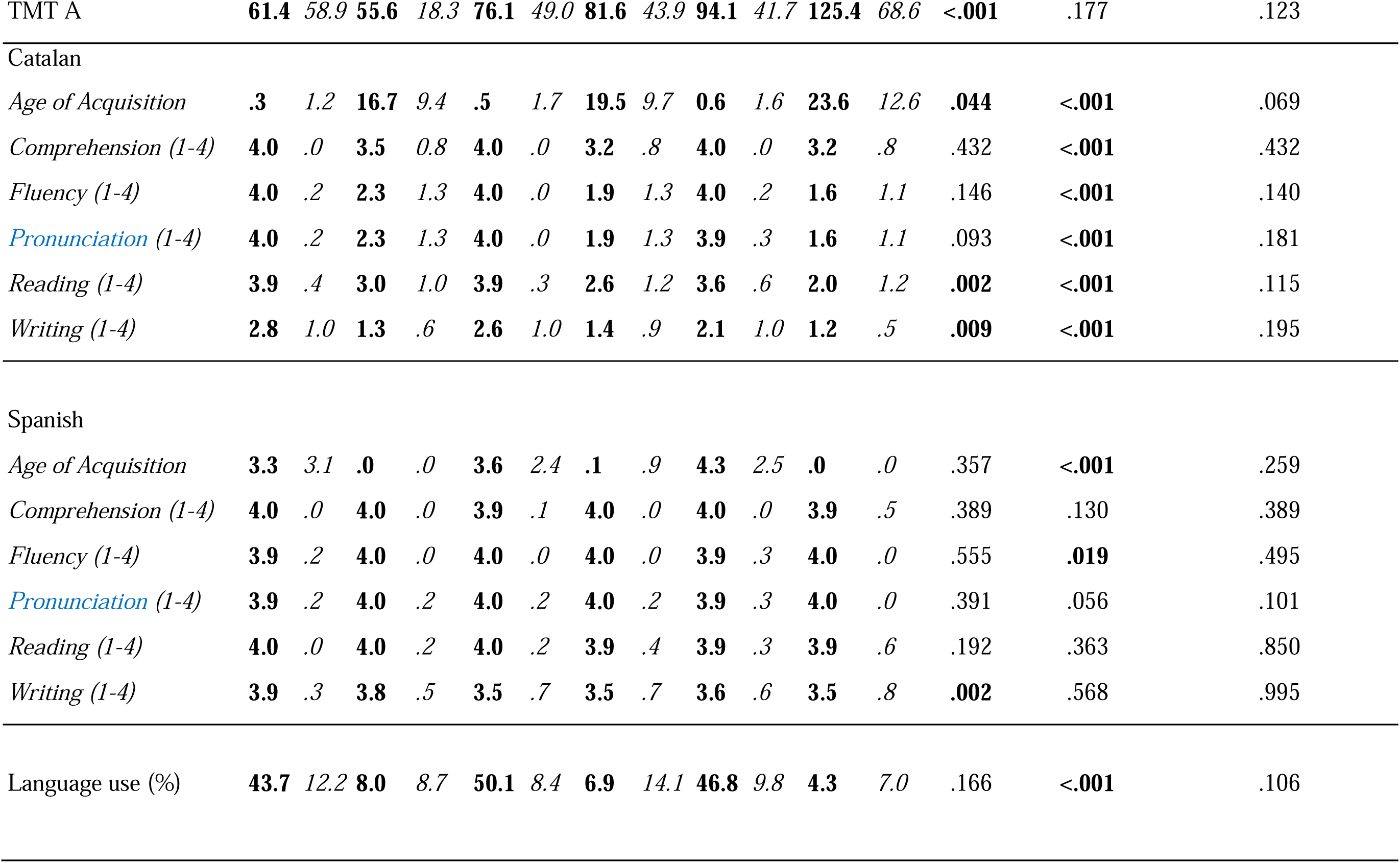
Mean values (M) and standard deviations (SD) for age, education, neuropsychological test score, and language profile, broken by group of participants (CU, MCI and AD) and Bilingualism type (active vs. passive). Significant p-values are marked in bold.

Clinical diagnoses of MCI and AD were established based on patients’ medical history, neurological examination, structural magnetic resonance imaging, and neuropsychological assessment, in accordance with established clinical criteria ^64,65^. Patients with potentially confounding neurological (other than MCI or AD) or psychiatric disorders, clinically known hearing or vision impairments, a past history of alcohol abuse, and/or psychosis, were excluded from the study. Healthy individuals had no previous neurological or psychiatric diseases. Additionally, healthy individuals were excluded from the study if they showed signs of cognitive deficits on a brief neuropsychological assessment (see Table 1).

### Linguistic profile

Language history and dominance were estimated using a questionnaire administered to participants. To enhance the accuracy of this information, we also conducted interviews with patients and some of their family members ^66^. In the current study, we excluded participants who spoke a third language to focus on bilingualism. The following measures were considered:

1. Age of acquisition of both languages (Catalan and Spanish). According to this variable, participants were classified as bilinguals if they had learned both languages before the age of 6 years.
2. Self-rating of their language proficiency consisted of their speaking fluency, comprehension, reading, and writing abilities on a four-point scale (1=poor, 2=regular, 3=good, 4=perfect).
3. Language usage was rated on a 0-100 scale. This measure represents the frequency with which participants used both languages in their daily lives from childhood to adulthood. If we express this variable in Catalan usage percentage, 0% will represent only Spanish use and 100% only Catalan use, while 50% will represent a balanced Catalan-Spanish usage. So, bilinguals were classified as such when their punctuation was between 40-60%.

As shown in Table 1, active bilinguals were those who had high proficiency in both Spanish and Catalan, spoke both languages in a balanced manner, and acquired both languages before the age of six. However, proficiency in writing and reading in Catalan was lower than in other linguistic domains within this group. This is due to older individuals being educated in Spanish, leading to a much lower self-rated proficiency in reading and writing in Catalan compared to Spanish. Although the age of Spanish acquisition and fluency in Spanish were significantly different from passive bilinguals, this group difference is not meaningful since their age of acquisition was below six and fluency was rated at 3.9 out of 4.

Passive bilinguals were Spanish-dominant, acquired Catalan significantly later and had lower speaking proficiency but higher comprehension in Catalan.

According to the questionnaire, 104 participants were classified as active bilinguals (CU= 32, MCI= 41, AD = 31) and 144 as passive bilinguals (CU = 26, MCI= 83, AD = 35).

### Neuropsychological measures

To assess participants, different hospital-specific neuropsychological batteries were applied in order to define participants’ cognitive decline degree, including the Mini-Mental State Examination (MMSE) which measures the degree of cognitive decline ^67^, forward and backward Digits Spans ^68^, which measure verbal short-term memory and working memory, the CERAD Word List Memory ^69^, which measures long-term episodic memory and the Trail Making Test part A ^68^ which measures visual attention and motor speed.

### Picture naming task

The materials consisted of 48 items representing different objects organised in 6 semantic categories: animals, vegetables, fruit, tools, furniture, and kitchen tools. We used black-and-white line drawings from Snodgrass and Vanderwart ^70^ as well as from the International Picture Naming Project Studies ^71^.

Word frequency and familiarity for Spanish and Catalan items were obtained from the NIM database ^72^. The items were selected across a range of word frequencies to test the effect of low and high frequency. The log word frequency for Spanish (M = 0.78, range = 0.13–1.69) and Catalan (M = 0.86, range = 0.10–1.74) did not differ significantly (p = .31). Familiarity ratings were also similar between Spanish (M = 6.07, range = 5.15–6.83) and Catalan (M = 6.42, range = 4.88–7.12), with no significant difference (p = .19). Mean word length, measured by number of letters, was not significantly different between Spanish (M = 6.39, range = 3–14) and Catalan (M = 6.04, range = 2–10; p = .42). However, the number of syllables was significantly different between Spanish (M = 2.81, range = 2–5) and Catalan (M = 2.42, range = 1–4; p < .05). This difference was minimal and likely reflects the tendency for Catalan words to be shorter than their Spanish counterparts.

Half the items for each frequency category were cognate words and half were non-cognate words. The word frequency for cognates and non-cognates was matched in Catalan (p = .597) and Spanish (p = .715).

Passive bilinguals performed the naming task in Spanish, whereas active bilinguals performed the task in Catalan and Spanish in two different sessions. However, for the purpose of this study, only the dominant language was compared between the two language groups. The naming language was always Spanish for the passive bilinguals and predominantly Catalan for the active bilinguals. Specifically, 94 active bilingual participants reported Catalan as their dominant language and were tested in Catalan (AD = 28, MCI = 38, CU = 28), whereas 10 reported Spanish as their dominant language and were tested in Spanish (AD = 3, MCI = 3, CU = 4). Language dominance in the active bilingual group was based on participants’ self-reported preference, as it could not be determined by age of acquisition or proficiency because both languages had been acquired simultaneously and were used with comparable proficiency.

Participants were asked to name the pictures that appeared on a laptop screen, with a maximum response time of 3.5 seconds. Pictures were preceded by a fixation point for 500 ms, and the intertrial interval was set at 1 second. All responses were recorded using DMDX ^73^ and analysed offline with Checkvocal ^74^.

### Data analyses

All statistical analyses were conducted using R (version 4.4.2).

Three types of outcomes were analyzed: accuracy (binary outcome), naming latencies (continuous outcome), and error types (counts). Naming latencies were log-transformed (logRT) to correct for non-normal distribution and skewness, as confirmed by visual inspection of histograms and Q-Q plots.

To analyse naming latencies and accuracy, mixed-effects models were fitted using the *lme4* package^75^. Log-transformed reaction times were analysed using linear mixed-effects models, while accuracy data were analysed using logistic mixed-effects models (binomial family).

Firstly, we compared naming accuracy and naming latencies between active and passive bilinguals in their dominant language. The initial models included fixed effects of Group (CU, MCI, and AD), log-transformed word frequency (Log Frequency), Cognate status (cognates vs. non-cognates), Bilingualism type (active vs. passive), and all interactions among these factors. CU served as the reference category for Group, cognates for Cognate status, and active bilinguals for Bilingualism type.

Random intercepts for participants and items were included in the accuracy models. The effect of word frequency was also tested as a categorical variable (high vs. low), instead of as a log-transformed continuous predictor.

We additionally ran extended models including standardized covariates (MMSE, age, and education). These covariates were included due to group differences in some of these variables and to control for individual variability in global cognitive status (MMSE), age, and years of education. Model selection was based on comparisons of the Akaike Information Criterion (AIC), Bayesian Information Criterion (BIC), and conditional and marginal R², using the *MuMIn* and *performance* packages.

Secondly, to further explore the effect of cognate status, we conducted separate analyses for cognate and non-cognate words. As this was an exploratory analysis, it may help to refine and improve the interpretation of the main analysis and was performed irrespective of the outcome of the primary model that included cognate status as a fixed effect. The fixed effects included Group (CU, MCI, and AD), log-transformed word frequency (Log Frequency), Bilingualism type (active vs. passive), and all interactions among these factors. CU served as the reference category for Group, cognates for Cognate status, and active bilinguals for Bilingualism type.

In addition, covariates that were significant in the extended model of the first analysis—where cognate and non-cognate words were analysed together—were included in these models.

All models were fitted using maximum likelihood estimation and the “bobyqa” optimizer (glmerControl(optimizer = “bobyqa”)) to ensure convergence for complex models. 95% Confidence Intervals (CIs) and p-values were computed using a Wald z-distribution approximation.

Finally, we compared the distribution of naming errors (i.e., the number of errors of each type) in both active and passive bilinguals with either MCI or AD. First, we classified the errors as follows: a) semantic: an incorrect response that was semantically related to the target, such as a category member or a superordinate (e.g., *squirrel* – *rabbit*; *apple* – *fruit*), or a circumlocution (i.e., a definition or description of the target is provided but the actual word is not produced); b) omission: when the participant was unable to name the item; c) cross-language intrusion: a correct word, but in the non-target language (e.g., for the item *rana* in Spanish, the participant said *granota* in Catalan, meaning ‘frog’ in English); d) visual: a response that was visually related to the target (e.g., *lettuce* – *rose*); e) unrelated: an incorrect response with no semantic or visual relationship to the target (e.g., *peach* – *ring*); f) mixed: a response that combined two of the error types above (e.g., for *cuc* in Catalan [‘worm’ in English], the participant said *serpiente* in Spanish [‘snake’ in English], which is both a semantic and a cross-language intrusion error); g) formal: a real-word response that differs from the target by one phoneme in a corresponding syllable or word position, or by two phonemes in any position^76^; and h) nonword: a string of phonemes that is not a real word in the language. Due to the low frequency of formal and nonword errors in our sample, both were grouped under a single category (other).

Subsequently, to analyse the distribution of errors across groups, a generalized linear mixed model was fitted using the *glmmTMB* package with a negative binomial distribution. The model included fixed effects of Group (MCI vs. AD), Bilingualism type (active vs. passive), Error Type (as classified above), their interactions, and covariates including education, age, and MMSE scores. CU served as the reference category for Group, cognates for Cognate status, active bilinguals for Bilingualism type, and omissions for Error Type. A random intercept for participants (1 | ID) was included to account for repeated measures within individuals.

This approach was chosen because the error data were count-based, sparse, and overdispersed. While a Poisson regression initially suggested overdispersion (dispersion > 1.5), the negative binomial distribution naturally accounts for this extra variability. Moreover, including random intercepts allows the model to capture individual differences and within-subject correlations, improving fit and generalizability. Importantly, this modelling approach does not rely on assumptions of normally distributed residuals, making it well suited for the observed distribution of errors.

Model assumptions were evaluated by inspecting residual distributions, leverage, and outliers with the *performance* and *see* packages. Multicollinearity was assessed using variance inflation factors (VIF) with the *car* package.

Categorical predictors were coded using treatment (dummy) coding (R default; *contr.treatment*), such that coefficients represent differences relative to a reference level. Continuous predictors were z-scored.

Model estimates and interactions were visualized using *ggplot2*, with predicted values generated from the models using the *ggeffects* and *effects* packages. Faceted plots by groups were generated where relevant, with stylistic enhancements to aid interpretation of group differences and interactions with bilingualism and word frequency.

Detailed results of the statistical models are provided in the Supplementary Materials, whereas only significant findings and final model outputs are reported in the Results section.

## Results

### Accuracy

The extended model, which included MMSE, age, and education as covariates, demonstrated a better fit, with lower AIC and BIC values (7643.1 and 7774.1, respectively) compared to the initial model without covariates (7906.5 and 8016.0). The proportion of variance explained by fixed effects alone (marginal R²) was 10%, while the proportion explained by both fixed and random effects (conditional R²) was 47% (see Table 2).

**Table 2.** Output model for accuracy with covariates: Accuracy ∼ Group * Log Frequency * Bilingualism type + Cognate + MMSE + Education + Age + (1 | ID) + (1 | ITEM)

| *Predictors* | **Accuracy** |  |  |
| --- | --- | --- | --- |
|  | *Estimates* | *CI* | *P* |
| (Intercept) | 3.09 | 2.53 – 3.66 | **<0.001** |
| Group [MCI] | -1.00 | -1.48 – -0.52 | **<0.001** |
| Group [AD] | -0.81 | -1.35 – -0.26 | **0.004** |
| Log Frequency | 0.32 | -0.04 – 0.68 | 0.081 |
| Bilingualism type [ Passive] | -0.36 | -0.96 – 0.24 | 0.236 |
| Cognate status [non-cognate] | -0.20 | -0.75 – 0.35 | 0.481 |
| MMSE | 0.21 | 0.04 – 0.38 | **0.015** |
| Age | -0.33 | -0.46 – -0.21 | **<0.001** |
| Education | 0.20 | 0.07 – 0.33 | **0.003** |
| Group [MCI] × Log Frequency | -0.08 | -0.34 – 0.18 | 0.558 |
| Group [AD] × Log Frequency | 0.06 | -0.21 – 0.33 | 0.679 |
| Group [MCI] × Bilingualism type [Passive] | 0.74 | 0.08 – 1.39 | **0.027** |
| Group [AD] × Bilingualism type [Passive] | -0.14 | -0.87 – 0.59 | 0.706 |
| Log Frequency × Bilingualism type [Passive] | -0.21 | -0.58 – 0.16 | 0.268 |
| Group [MCI] × Log Frequency × Bilingualism type [Passive] | 0.18 | -0.17 – 0.54 | 0.311 |
| Group [AD] × Log Frequency × Bilingualism type [Passive] | 0.27 | -0.11 – 0.64 | 0.162 |
| AIC/BIC | 7643.1/7774.1 |  |  |
| Marginal R<sup>2</sup> / Conditional R<sup>2</sup> | 0.097 / 0.468 |  |  |

Diagnostics for residuals and outliers did not reveal any significant issues. The Kolmogorov–Smirnov test for residual distribution showed p = 0.803, indicating no significant deviation from the expected distribution. The dispersion test returned p = 0.864, showing no evidence of overdispersion or underdispersion. The outlier test resulted in p = 0.252, suggesting no influential outliers distorting the results. DHARMa simulated residual plots confirmed no deviations between predicted and simulated residuals. None of the predictors or interaction terms showed VIF values above 5, indicating no multicollinearity among predictors.

The effect of Group was statistically significant, meaning that MCI (β = −1.01, 95% CI [−1.48, −0.52], p < .001) and AD patients (β = −0.81, 95% CI [−1.35, −0.26], p = .004) named fewer pictures than CU.

The interaction between Group (MCI) and Bilingualism Type (passive) was statistically significant and positive (β = 0.74, 95% CI [0.08, 1.39], p = .027), indicating that passive bilinguals with MCI named more pictures than their monolingual counterparts (see Figure 1).

**FIGURE 1.**
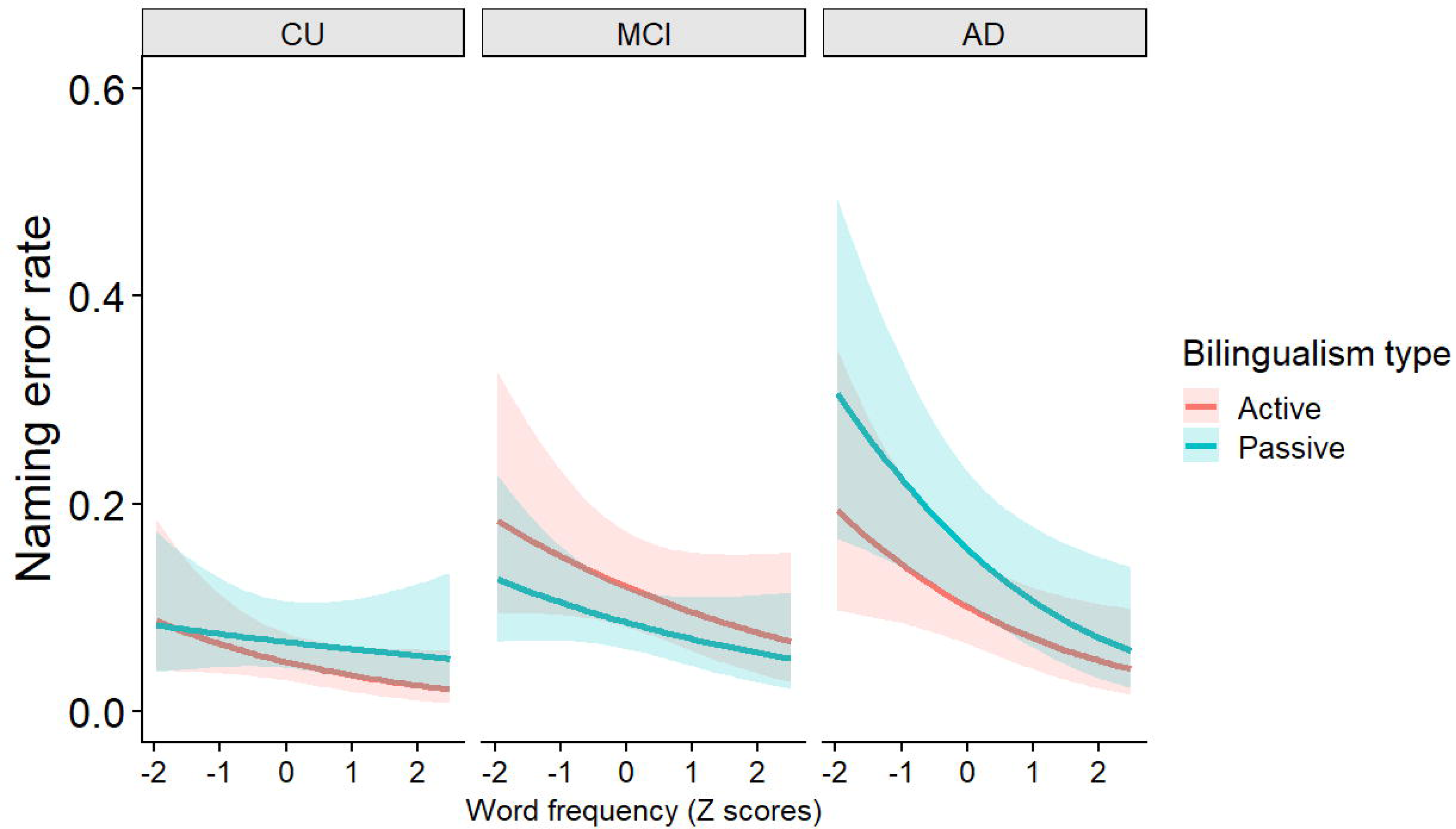

We further analysed the data using word frequency as a categorical variable (high vs. low). The results of this model were comparable to those obtained with log-transformed frequency (for more details, see the section “Accuracy: categorical frequency effect” in the Supplementary Materials).

Additionally, we reanalysed the data by separating cognates and non-cognates (for more details, see the section “Accuracy: cognate status effect” in the Supplementary Materials).

For <u>non-cognates</u>, participants with MCI (β = −1.17, 95% CI [−1.75, −0.60], p < .001) and AD (β = −1.20, 95% CI [−1.83, −0.57], p < .001) named fewer pictures than CU. Additionally, the interaction between Group (MCI) and Bilingualism Type was significant (β = 0.85, 95% CI [0.08, 1.62], p = .031), indicating that passive bilinguals named more pictures than active bilinguals.

For cognates, participants with MCI were less accurate than CU (β = −0.81, 95% CI [−1.33, −0.29], p = .002).

### Naming latencies

The extended model showed a better fit with lower AIC and BIC values (5809.6/ 5937.2) than the initial model without covariates (5922.9/ 6036.6). The proportion of variance explained by fixed effects alone (marginal R²) was 9%, while the proportion explained by both fixed and random effects (conditional R²) was 32%. 95% CIs and p-values were computed using a Wald z-distribution approximation (see Table 3).

**Table 3.** Output model for naming latencies with covariates: Log RT ∼ Group * Log Frequency * Bilingualism type + Cognate + MMSE+ Education + (1 | ID) + (1 | ITEM)

| *Predictors* | **log(RT)** |  |  |
| --- | --- | --- | --- |
|  | *Estimates* | *CI* | *p* |
| (Intercept) | 7.22 | 7.15 – 7.28 | **<0.001** |
| Group [MCI] | 0.06 | -0.01 – 0.12 | 0.106 |
| Group [AD] | 0.15 | 0.08 – 0.23 | **<0.001** |
| Log Frequency | -0.02 | -0.05 – 0.01 | 0.229 |
| Bilingualism type [Passive] | 0.05 | -0.03 – 0.13 | 0.194 |
| Cognate status [non-cognate] | 0.02 | -0.04 – 0.08 | 0.513 |
| MMSE | -0.03 | -0.06 – -0.01 | **0.007** |
| Age | 0.03 | 0.01 – 0.05 | **0.001** |
| Group [MCI] × Log Frequency | -0.01 | -0.03 – 0.02 | 0.534 |
| Group [AD] × Log Frequency | 0.01 | -0.01 – 0.04 | 0.371 |
| Group [MCI] × Bilingualism type [Passive] | -0.01 | -0.10 – 0.08 | 0.883 |
| GROUP [AD] × Bilingualism type [Passive] | 0.06 | -0.05 – 0.16 | 0.291 |
| Log Frequency × Bilingualism type [Passive] | -0.03 | -0.06 – -0.01 | **0.044** |
| Group [MCI] × Log Frequency × Bilingualism type[Passive] | 0.00 | -0.03 – 0.04 | 0.887 |
| Group [AD] × Log Frequency × Bilingualism type [Passive] | 0.04 | 0.01 – 0.08 | **0.041** |
| AIC/BIC | 5809.6/ 5937.2 |  |  |
| Marginal R<sup>2</sup> / Conditional R<sup>2</sup> | 0.087 / 0.323 |  |  |

The effect of Group was significant for the AD group compared with CU (β = 0.15, 95% CI [0.08, 0.23], p < .001), suggesting that individuals with AD were slower in naming compared to CU. Conversely, the MCI group performed similarly to CU (β = 0.06, 95% CI [−0.01, 0.12], p = .106).

The interaction between Log frequency and Bilingualism type (β = −0.03, 95% CI [−0.06, −0.01], p = .044), as well as the three-way interaction between Log frequency, Group (AD), and Bilingualism type (β = 0.04, 95% CI [0.01, 0.08], p = .041), were significant. This pattern suggests that the effect of word frequency was larger in passive bilinguals, although this pattern reversed in patients with AD (see Figure 2).

**FIGURE 2.**
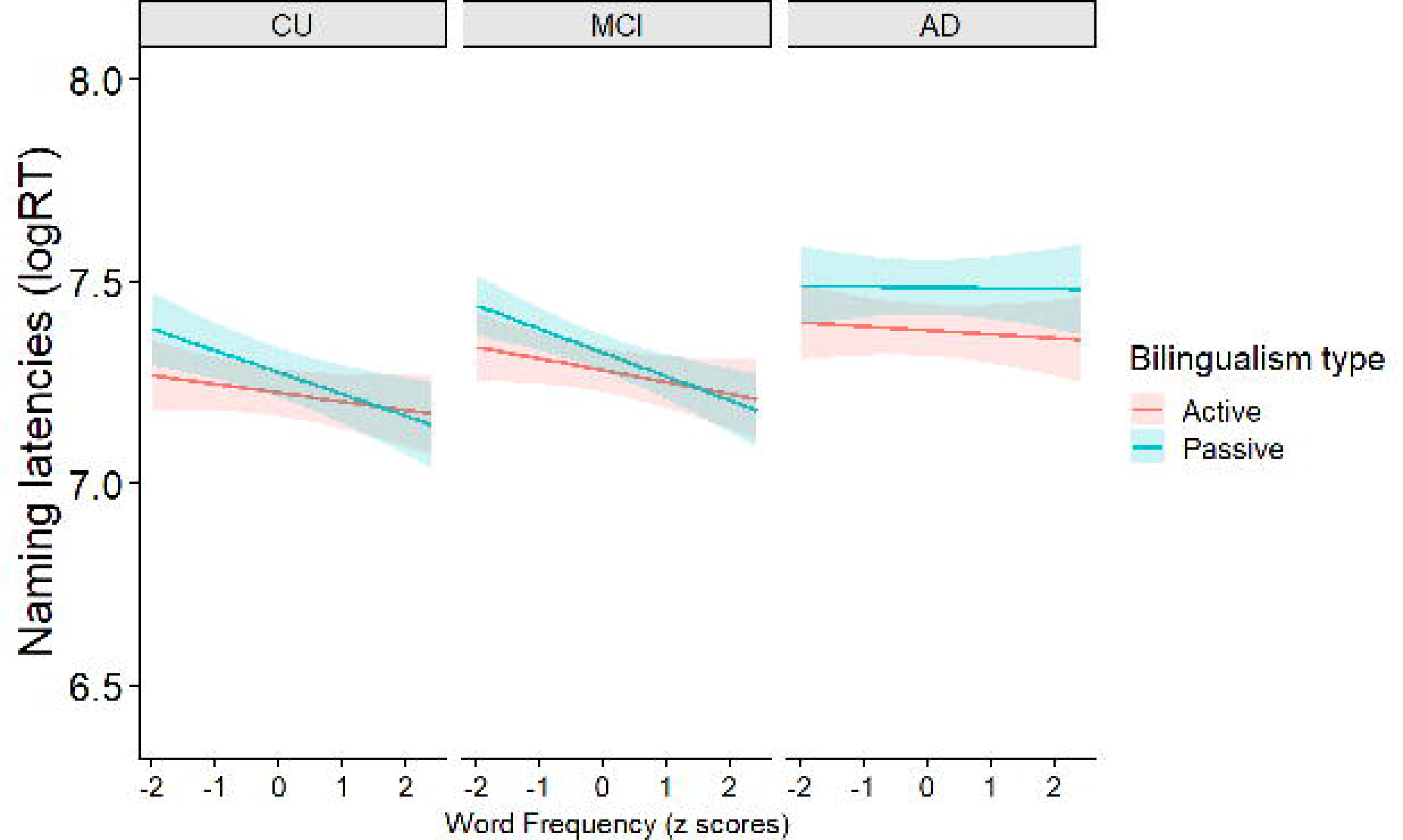

We further analysed the data using word frequency as a categorical variable (high vs. low) and the results of this model were comparable to those obtained with log-transformed frequency (for more details, see the section “Naming latencies: categorical frequency effect” in the Supplementary Materials).

Additionally, we reanalysed the data by separating cognates and non-cognates (for more details, see the section “Naming latencies: cognate status effect” in the Supplementary Materials).

For <u>non-cognates</u>, the effect of Group was significant for the AD group compared with CU (β = 0.14, 95% CI [0.05, 0.22], p = .001), suggesting that individuals with AD were slower in naming compared to CU. Additionally, the effect of Log frequency was significant (β = −0.06, 95% CI [−0.11, −0.01], p = .033), indicating faster naming latencies for high-frequency words than low-frequency words.

For cognates, the effect of Group was significant for both the AD group (β = 0.19, 95% CI [0.11, 0.28], p < .001) and the MCI group (β = 0.08, 95% CI [0.01, 0.15], p = .026), compared with CU, suggesting that individuals with AD and MCI were slower in naming compared to CU.

### Error types

The extended model demonstrated superior fit, as indicated by a lower AIC and BIC value (3052.7/3223.2) compared to the initial model without covariates (3169.9/3325.6) (See Table 4).

**Table 4.** Output model for error type with covariates: Errors ∼ Error Type * Group * Bilingualism type + Education+ Age + MMSE + (1 | ID)

| *Predictors* | **Errors** |  |  |
| --- | --- | --- | --- |
|  | *Estimates* | *CI* | *p* |
| (Intercept) | 1.08 | 0.80 – 1.37 | **<0.001** |
| Error Type [Semantic] | -0.02 | -0.40 – 0.36 | 0.919 |
| Error Type [CL Intrusion] | -0.30 | -0.70 – 0.10 | 0.143 |
| Error Type [Visual] | -4.11 | -5.53 – -2.68 | **<0.001** |
| Error Type [Unrelated] | -2.60 | -3.34 – -1.87 | **<0.001** |
| Error Type [Mixed] | -1.64 | -2.16 – -1.11 | **<0.001** |
| Error Type [Other] | -3.69 | -4.87 – -2.51 | **<0.001** |
| Group [AD] | 0.24 | -0.16 – 0.64 | 0.246 |
| Bilingualism type [Passive] | -0.59 | -0.95 – -0.23 | **0.001** |
| Education | -0.10 | -0.19 – -0.01 | **0.022** |
| Age | 0.20 | 0.12 – 0.28 | **<0.001** |
| MMSE | -0.14 | -0.23 – -0.04 | **0.004** |
| Error Type [Semantic] × Group [AD] | -0.09 | -0.64 – 0.46 | 0.743 |
| Error Type [CL Intrusion] × Group [AD] | -1.21 | -1.85 – -0.57 | **<0.001** |
| Error Type [Visual] × Group [AD] | 0.52 | -1.25 – 2.28 | 0.568 |
| Error Type [Unrelated] × Group [AD] | -0.56 | -1.71 – 0.59 | 0.338 |
| Error Type [Mixed] × Group [AD] | -1.55 | -2.58 – -0.53 | **0.003** |
| Error Type [Other] × Group [AD] | -0.60 | -2.45 – 1.26 | 0.528 |
| Error Type [Semantic] × Bilingualism type [Passive] | 0.89 | 0.42 – 1.37 | **<0.001** |
| Error Type [CL Intrusion] × Bilingualism type [Passive] | -0.72 | -1.27 – -0.16 | **0.011** |
| Error Type [Visual] × Bilingualism type [Passive] | 2.38 | 0.87 – 3.88 | **0.002** |
| Error Type [Unrelated] × Bilingualism type [Passive] | 1.31 | 0.46 – 2.15 | **0.003** |
| Error Type [Mixed] × Bilingualism type [Passive] | -0.74 | -1.55 – 0.08 | 0.075 |
| Error Type [Other] × Bilingualism type [Passive] | 1.03 | -0.35 – 2.40 | 0.143 |
| Group [AD] × Bilingualism type [Passive] | 0.54 | 0.01 – 1.07 | **0.045** |
| Error Type [Semantic] × Group [AD] × Bilingualism type [Passive] | -0.32 | -1.03 – 0.40 | 0.388 |
| Error Type [CL Intrusion] × Group [AD] × Bilingualism type [Passive] | 0.57 | -0.33 – 1.47 | 0.213 |
| Error Type [Visual] × Group [AD] × Bilingualism type [Passive] | -0.86 | -2.77 – 1.06 | 0.380 |
| Error Type [Unrelated] × Group [AD] × Bilingualism type [Passive] | -0.32 | -1.67 – 1.03 | 0.640 |
| Error Type [Mixed] × Group [AD] × Bilingualism type [Passive] | 0.65 | -0.83 – 2.13 | 0.392 |
| Error Type [Other] × Group [AD] × Bilingualism type [Passive] | -0.72 | -3.03 – 1.59 | 0.540 |
| AIC/BIC | 3052.3223.2 |  |  |
| Marginal R<sup>2</sup> / Conditional R<sup>2</sup> | 0.716 / 0.740 |  |  |

Relative to the reference category (omission errors), semantic errors (β = −0.02, 95% CI [−0.40, 0.36], p = .92) and cross-language intrusion errors (β = −0.30, 95% CI [−0.70, 0.10], p = .14) did not differ significantly, indicating comparable frequencies of occurrence.

In contrast, unrelated (β = −2.60, 95% CI [−3.34, −1.87], p < .001), visual (β = −4.11, 95% CI [−5.53, −2.68], p < .001), mixed (β = −1.64, 95% CI [−2.16, −1.11], p < .001), and other errors (β = −3.69, 95% CI [−4.87, −2.51], p < .001) occurred significantly less frequently than omission errors (see Figure 3).

**FIGURE 3.**
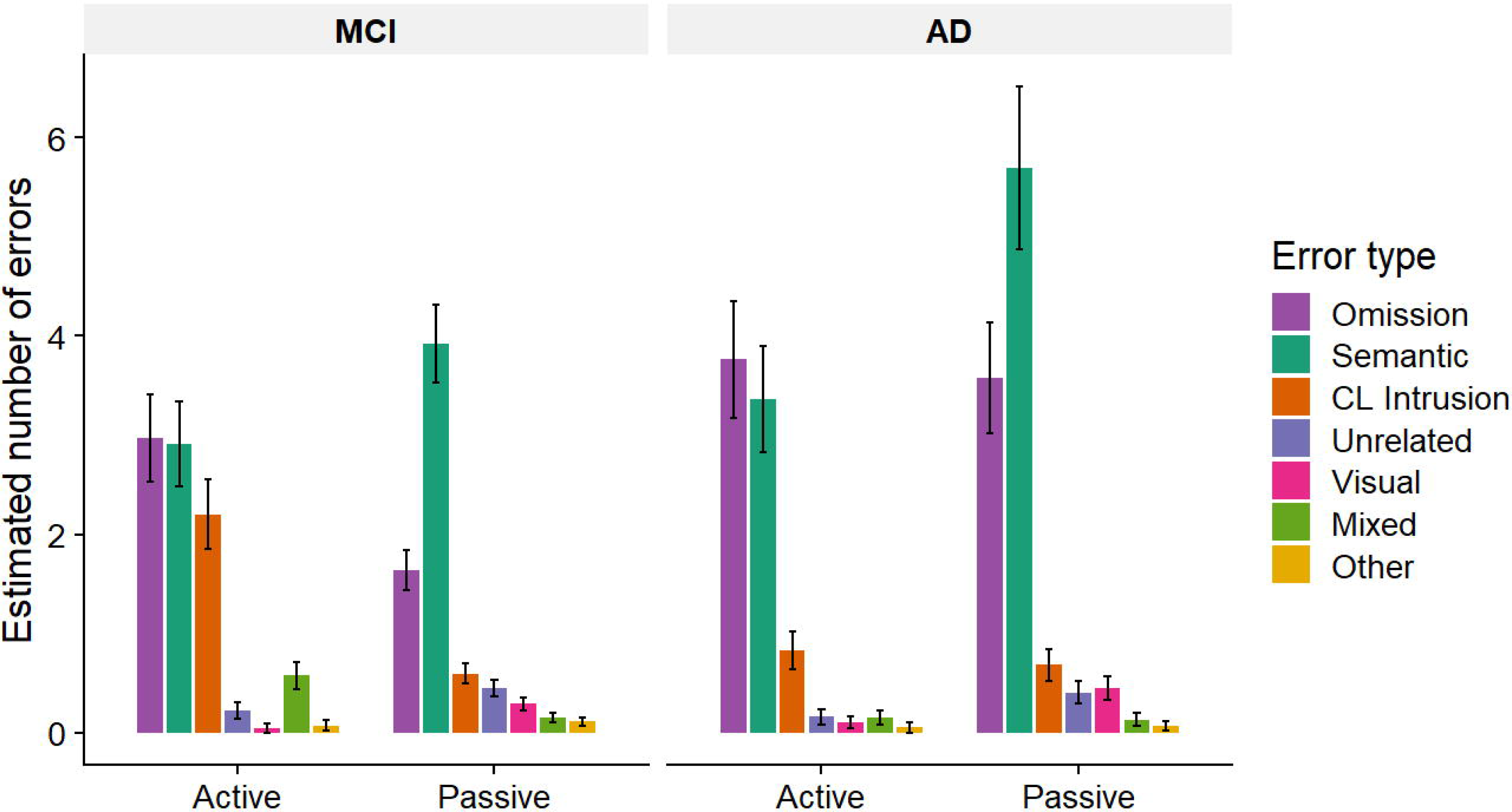

The effect of Bilingualism type (passive) was negative and significant (β = −0.59, 95% CI [−0.95, −0.23], p = .001), indicating that passive bilinguals with MCI produced fewer errors overall than active bilinguals.

Several significant interactions were identified between error type and both Group and Bilingualism type.

First, the interaction between cross-language intrusions and Group (AD) was negative and significant (β = −.25, 95% CI [-.85, −0.57], p < .001), indicating that AD patients produced fewer cross-language intrusions compared to participants with MCI. Second, the interaction between cross-language intrusions and Bilingualism type (passive) was negative and significant (β = −0.72, 95% CI [−1.27, −0.16], p = .011), indicating that cross-language intrusions were less frequent among passive bilinguals.

Third, the interactions between semantic errors and Bilingualism type (passive) (β = 0.89, 95% CI [0.42, 1.37], p < .001), visual errors and bilingualism type (passive) (β = 2.38, 95% CI [0.87, 3.88], p = .002), and unrelated errors and Bilingualism type (passive) (β = 1.31, 95% CI [0.46, 2.15], p = .003) were positive, suggesting that these error types were more frequent among passive bilinguals.

Finally, a significant interaction between Group and Bilingualism type (passive) was observed (β = 0.54, 95% CI [0.01, 1.07], p = .045), indicating that passive bilinguals with AD produced more errors than active bilinguals.

## Discussion

In this study, we examined the extent to which active bilingualism—characterized by high proficiency and balanced use of two languages—modulates lexical access in the dominant language of patients with MCI and AD, compared with CU. Although a growing body of research has focused on healthy bilingual populations, studies in the context of neurodegenerative diseases remain limited. To address this gap, we focused on analysing naming performance by distinguishing between two types of bilingualism, rather than contrasting bilinguals with monolinguals, to better reflect the sociolinguistic context of the speakers.

Compared to cognitively unimpaired adults, both patient groups (MCI and AD) exhibited significant word-finding deficits in the naming task. This finding aligns with previous research showing that naming deficits emerge from the preclinical stage of AD, as observed in our MCI patients ^2,60,61,77^. Although MCI patients did not show significantly slower naming latencies, they exhibited mild word-finding deficits in accuracy. In both MCI and AD groups, semantic and omission errors were the most frequent, consistent with previous research in naming tasks.

Based on the previous findings of a bilingual disadvantage in word production tasks among CU older adults ^14,16,28,29^, our main hypothesis predicted that active bilingual speakers would exhibit greater word-finding difficulties than passive bilinguals. In our sample, passive bilinguals were Spanish-dominant speakers with passive exposure to Catalan but limited spoken proficiency in this language. Therefore, they were expected to perform the naming task similarly to monolinguals, who predominantly or exclusively use only one language in their daily life. This prediction aligns with the frequency-lag hypothesis^26^ which posits a direct relationship between the frequency of language use and the bilingual disadvantage.

Regarding bilingual status, CU older adults from both language groups showed comparable naming accuracy. However, active bilinguals tended to outperform passive bilinguals in naming latencies; specifically, they were faster at naming at naming low-frequency pictures. This initial pattern was confirmed in the general analysis of naming latencies for MCI, but not for AD who were non sensitive to the interaction between word frequency and naming latencies. The further analysis conducted by separating cognates and non-cognates words reveal a more complex pattern of results. Specifically, for cognate words—those that share orthographic and phonological features across languages—we observed a trend toward faster responses in active bilinguals compared to passive bilinguals. Although the difference between the two language groups was small, it was consistent across diagnostic groups. It is important to note that this difference reached only marginal significance, so caution is warranted when interpreting these results.

This potential advantage of active bilingualism can be explained by the benefits of having shared morphological, lexical, or phonological representations for these types of words (for a discussion, see ^56^). Specifically, in our sample, the benefits likely arise from speaking a second language with a high frequency of cognates, as is the case for our Catalan-Spanish bilinguals. Consistently, in our previous studies examining patterns of decline in Catalan-Spanish bilinguals, cognates were generally less affected by neurodegenerative diseases than non-cognate words ^43,44^.

To summarize, these naming latency results suggest that active bilinguals retain a certain degree of lexical retrieval efficiency, both in cognitively unimpaired individuals and in those at the preclinical stage of AD, such as MCI.

Besides this possible advantage in lexical processing efficiency, we also considered the results for naming accuracy, which help determine whether this benefit extends to word-finding abilities, such as lexical selection and retrieval. It is well established that individuals with MCI and AD may exhibit word-finding deficits due to multiple factors, including impairments in lexical access, access to the phonological lexicon, and/or semantic processing^40–42^. In our sample, we found evidence of such naming deficits, with a trend toward modulation by word frequency, consistent with previous findings showing that low-frequency words are disproportionately impaired in these patients compared to CU ^54,55^.

With respect to bilingual status, we did not observe an overall difference in performance across groups. However, for non-cognates, individuals with MCI in the active bilingual group consistently produced more errors than those in the passive bilingual group. This pattern was not observed for cognates, where the effect was reduced and not statistically significant.

This pattern aligns with previous studies reporting better naming performance in monolinguals than in bilinguals and is consistent with the frequency-lag hypothesis ^26^. For instance, Gard et al.^46^ reported superior performance in monolinguals with MCI on the Boston Naming Test, though not on verbal fluency measures, while Anderson et al.^28^ found that bilinguals with AD (but not MCI) showed significantly poorer performance on letter fluency tasks.

Thus, there appears to be a relative impairment in retrieving words, alongside faster processing for words unaffected in the preclinical stages of the disease—likely due to preserved phonological processing, which provides facilitation. This pattern could have reflected a speed–accuracy trade-off between naming latencies and accuracy. However, we tested for a correlation between these two measures and found a negative correlation in all participant groups, allowing us to rule out this interpretation (see Supplementary Material).

To further elucidate the nature of these naming deficits, we examined the patterns of error types produced by the patient groups. We acknowledge that the low frequency of certain error types may reduce the statistical power to detect significant effects; therefore, we focus our discussion on the three most frequent error categories in our sample: omissions, semantic errors, and cross-language intrusions.

In patients with MCI, the active bilingual group produced more omission errors than the passive bilingual group, whereas the opposite pattern was observed for semantic errors. This increase in semantic errors became more pronounced in the AD groups, resulting in a higher overall error rate among passive bilinguals. This pattern aligns with the more general advantage in processing speed observed in the analysis of naming latencies and was only marginally significant for cognates in the naming accuracy analysis.

We propose that the advantage observed in active bilinguals with AD may reflect a relative preservation of semantic memory. In MCI, this pattern is less evident, likely due to the relatively mild semantic impairment at this stage, but it becomes more apparent in the AD group as semantic deficits progress. Although this interpretation is based primarily on error-type analyses, it is consistent with previous research attributing superior naming performance in bilinguals to better preserved or more robust semantic memory networks. For example, Arce et al. ^78^ reported that active Spanish–English bilinguals outperformed monolinguals in semantic, but not episodic, memory, and that active bilingualism was associated with protection against cortical thinning in the entorhinal cortex. Similarly, Taler and Johns ^31^ found that English–French bilinguals outperformed English monolinguals on a verbal fluency task, an effect they attributed to more efficient access to semantically distant words within the semantic network. This interpretation aligns with the semantic diversity account, which has also been proposed to explain bilingual advantages in older adults and differences in speech production performance between bilinguals and monolinguals ^79^.

However, there remains to be explained the origin of the worse performance of the active bilingual group with MCI compared to the passive bilingual group. Part of this results might be explained by more omissions, which indicates a failure to retrieve the correct word.

In addition, active bilinguals with MCI produced more cross-language intrusions than passive bilinguals, retrieving the correct target but in the non-target language. Although the overall frequency of this error type was low and caution is therefore warranted, we may speculate that this increase could reflect two main mechanisms. First, cross-language intrusions may constitute a compensatory strategy^63^ to facilitate communication when lexical retrieval difficulties arise, particularly in the sociolinguistic context of our participants, where code-switching is generally considered appropriate and meaningful by speakers ^80^.

However, if cross-language intrusions were primarily driven by deficits in language control, their frequency would be expected to increase at the AD stage, given the greater overall cognitive impairment in these patients. This pattern was not observed in our data; instead, omissions and semantic errors were the types of errors that tended to increase from MCI to AD. Furthermore, we examined whether cross-language intrusions were more likely to occur for cognates, as previous studies have shown that under certain naming conditions, cognates may induce interference rather than facilitation. Only 32% of cross-language intrusions involved cognates, a proportion lower than that observed for other common error types. Therefore, although we cannot rule out the possibility that the increased number of cross-language intrusions in active bilinguals with MCI reflects deficits in language control, we consider this explanation less likely.

Despite the relevance of examining how bilingualism modulates lexical access in patients with MCI and AD, we acknowledge that our findings are limited by the characteristics of our bilingual sample. Specifically, we compared active and passive bilinguals rather than monolinguals (i.e., individuals with no exposure to a second language), which may have influenced the observed effects. Although this limitation could be considered a weakness, we argue that focusing on this specific bilingual distinction provides valuable insight into how different sociolinguistic contexts modulate language deterioration in individuals with neurodegenerative diseases. Interestingly, we observed a delayed age at diagnosis and symptom onset in active bilinguals compared to passive bilinguals, consistent with our previous findings using a continuous measure of bilingualism ^81^. This observation opens new avenues for research, particularly in other bilingual communities with speakers exhibiting varying degrees of language experience.

We acknowledge that our findings should be interpreted considering several limitations of our study.

First, our bilingual participants differed in terms of education level and global cognitive scores. To account for this, we included MMSE scores and years of education as covariates in the model; the results remained unchanged. Therefore, although we cannot entirely rule out the possibility that passive bilinguals had greater disease severity or lower education affecting their performance, we are confident that the naming advantages observed in active bilinguals relative to passive bilinguals cannot be fully explained by these factors.

Second, the classification of our participants as active or passive bilinguals relied on subjective measures of language proficiency. Consequently, disease progression may have caused some patients—particularly those in the AD stage—to become more dominant in one language, potentially leading to misclassification. The inclusion of objective measures, such as naming performance ^33^ or vocabulary knowledge in both languages, would have strengthened the classification of our bilingual participants.

In addition, the two languages spoken by the active bilingual participants were Catalan and Spanish, two Romance languages that are typologically similar, particularly at the lexical level. This linguistic proximity may have influenced our findings, besides having analysed cognates and non-cognates separately. Indeed, in a previous study of Catalan–Spanish bilinguals with MCI, we observed a slower rate of decline in verbal fluency compared to Spanish monolinguals, supporting the potential role of language similarity in modulating the linguistic outcomes ^49^. While this is a plausible explanation—given reviews highlighting the influence of language distance on the engagement of different brain areas within the bilingual language network ^82^ and on patterns of recovery in aphasia ^83^—its role remains unclear, as other reviews suggest that language distance does not have a major impact on naming deficits ^84^.

In conclusion, our findings indicate that neurocognitive disorders such as MCI and AD may affect bilingual speech production in multiple ways. Active bilingualism appears to confer an advantage in lexical retrieval efficiency in terms of processing speed, likely due to better-preserved phonological processing that maintains facilitation^57^. However, at the MCI stage, active bilinguals seem to experience greater anomia, probably reflecting lexical or morphological retrieval deficits. In later stages, when the semantic network is more impaired, speaking a second language may provide an advantage by enriching semantic representations, particularly in advanced disease stages. These results should be interpreted with caution, as some effects may be influenced by difficulties in controlling the two languages.

Further research is needed to examine the role of different types of bilingualism across a wider range of language pairs and bilingual communities, to deepen our understanding of how language experience shapes lexical retrieval deficits. Such studies would not only advance theoretical accounts of bilingual language production but also have important practical implications for the assessment of language abilities in bilingual individuals with neurodegenerative diseases.

## Acknowledgements

We would like to thank all participants for their time and collaboration. We also thank the two anonymous reviewers, whose insightful comments helped improve both our manuscript and the interpretation of our findings.

## Author contributions

<u>María Sainz-Pardo</u>: Conceptualization, Formal analysis, Investigation, Data Curation, Writing - Original Draft, Writing - Review & Editing

<u>Mireia Hernández</u>: Methodology, Investigation, Writing - Original Draft, Writing - Review & Editing

<u>Anna Suades</u>: Methodology, Investigation, Resources, Writing - Original Draft, Writing - Review & Editing

<u>Montserrat Juncadella</u>: Methodology, Investigation, Resources, Writing - Original Draft, Writing - Review & Editing

<u>Jordi Ortiz-Gil</u>: Methodology, Investigation, Resources, Writing - Original Draft, Writing - Review & Editing

<u>Lidia Ugas</u>: Methodology, Investigation, Resources,Writing - Original Draft, Writing - Review & Editing

<u>Isabel Sala</u>: Methodology, Investigation, Resources, Writing - Original Draft, Writing - Review & Editing

<u>Alberto Lleó</u>: Methodology, Investigation, Resources, Writing - Original Draft, Writing - Review & Editing

<u>Marco Calabria</u>: Conceptualization, Methodology, Formal analysis, Investigation, Funding acquisition, Writing - Original Draft, Writing - Review & Editing

## Statements and declarations

Not applicable

## Ethical considerations

This investigation was carried out under the approval of ‘Parc de Salut MAR’ Research Ethics Committee under the reference number 2014/6003/I.

## Consent to participate

All participants signed an informed consent form, were fully informed about the study procedures, and understood that their participation was entirely voluntary. The consent form and study procedures were approved by ‘Parc de Salut MAR’ Research Ethics Committee under the reference number 2014/6003/I.

## Declaration of conflicting interest

All authors disclose no actual or potential conflicts of interest, including any financial, personal, or other relationships with individuals or organizations that could inappropriately influence (bias) their work.

Marco Calabria serves as Associate Editor for the Journal of Alzheimer’s Disease.

## Data availability

The raw data for the naming analysis are available at https://osf.io/4m3uj/files

## Artificial Intelligence Use Statement

The authors declare that artificial intelligence (AI) tools were used solely for proofreading and improving the clarity of the English language in this manuscript. No AI tools were used for data analysis, interpretation, or generation of scientific content. All scientific content, interpretations, and conclusions are the sole responsibility of the authors.

## References

1. Laws KR, Adlington RL, Gale TM, et al. A meta-analytic review of category naming in Alzheimer’s disease. Neuropsychologia 2007; 45: 2674–2682.

2. Taler V, Phillips NA. Language performance in Alzheimer’s disease and mild cognitive impairment: A comparative review. J Clin Exp Neuropsychol 2008; 30: 501–556.

3. Verma M, Howard RJ. Semantic memory and language dysfunction in early Alzheimer’s disease: a review. Int J Geriatr Psychiatry 2012; 27: 1209–1217.

4. Kavé G, Goral M. Do age-related word retrieval difficulties appear (or disappear) in connected speech? *Aging*, Neuropsychol Cogn 2017; 24: 508–527.

5. Stilwell BL, Dow RM, Lamers C, et al. Language changes in bilingual individuals with Alzheimer’s disease. International Journal of Language and Communication Disorders 2016; 51: 113–127.

6. Costa A, Santesteban M, Caño A. On the facilitatory effects of cognate words in bilingual speech production. Brain Lang 2005; 94: 94–103.

7. Kroll JF, Bobb SC, Misra M, et al. Language selection in bilingual speech: Evidence for inhibitory processes. Acta Psychol (Amst) 2008; 128: 416–430.

8. Kroll JF, Bobb SC, Wodniecka Z. Language selectivity is the exception, not the rule: Arguments against a fixed locus of language selection in bilingual speech. Biling Lang Cogn 2006; 9: 119–135.

9. La Heij W. Selection processes in monolingual and bilingual lexical access. In: Handbook of Bilingualism Psycholinguistic Approaches. 2005, pp. 289–307.

10. Gollan TH, Montoya RI, Cera C, et al. More use almost always a means a smaller frequency effect: Aging, bilingualism, and the weaker links hypothesis. J Mem Lang 2008; 58: 787–814.

11. Gollan TH, Acenas L-AR. What is a TOT? Cognate and translation effects on tip-of-the-tongue states in Spanish-English and tagalog-English bilinguals. J Exp Psychol Learn Mem Cogn 2004; 30: 246–269.

12. Basnight-Brown DM. Models of lexical access and bilingualism. In: Foundations of Bilingual Memory. 2014. Epub ahead of print 2014. DOI: 10.1007/978-1-4614-9218-4_5.

13. Gollan TH, Montoya RI, Fennema-Notestine C, et al. Bilingualism affects picture naming but not picture classification. Mem Cognit 2005; 33: 1220–1234.

14. Bialystok E. Effects of bilingualism on cognitive and linguistic performance across the lifespan. In: Streitfall Zweisprachigkeit–The Bilingualism Controversy. Springer, 2009, pp. 53–67.

15. Bialystok E, Craik FIM. Cognitive and linguistic processing in the bilingual mind. Curr Dir Psychol Sci 2010; 19: 19–23.

16. Rosselli M, Ardila A, Araujo K, et al. Verbal fluency and repetition skills in healthy older Spanish-English bilinguals. Appl Neuropsychol 2000; 7: 17–24.

17. Tahami Monfared AA, Byrnes MJ, White LA, et al. Alzheimer’s disease: epidemiology and clinical progression. Neurol Ther 2022; 11: 553–569.

18. García AM, de Leon J, Tee BL, et al. Speech and language markers of neurodegeneration: a call for global equity. Brain 2023; 146: 4870–4879.

19. Bylund E, Antfolk J, Abrahamsson N, et al. Does bilingualism come with linguistic costs? A meta-analytic review of the bilingual lexical deficit. Psychon Bull Rev 2023; 30: 897–913.

20. Bailey C, Venta A, Langley H. The bilingual [dis] advantage. Lang Cogn 2020; 12: 225–281.

21. Gollan TH, Kroll JF. Bilingual lexical access. In: Handbook of Cognitive Neuropsychology. Psychology Press, 2015, pp. 321–345.

22. Green DW. Mental control of the bilingual lexico-semantic system. Biling Lang Cogn 1998; 1: 67–81.

23. Santesteban M, Schwieter JW. Lexical selection and competition in bilinguals. Biling Lex ambiguity Resolut 2020; 126–156.

24. Colomé À. Lexical activation in bilinguals’ speech production: Language-specific or language-independent? J Mem Lang 2001; 45: 721–736.

25. Kroll JF, Ma F. The Bilingual Lexicon. In: The Handbook of Psycholinguistics, pp. 294–319.

26. Gollan TH, Slattery TJ, Goldenberg D, et al. Frequency drives lexical access in reading but not in speaking: the frequency-lag hypothesis. J Exp Psychol Gen 2011; 140: 186.

27. Ivanova I, Costa A. Does bilingualism hamper lexical access in speech production? Acta Psychol (Amst) 2008; 127: 277–288.

28. Anderson JAE, Saleemi S, Bialystok E. Neuropsychological Assessments of Cognitive Aging in Monolingual and Bilingual Older Adults. J Neurolinguistics 2017; 43: 17–27.

29. Sullivan MD, Poarch GJ, Bialystok E. Why is lexical retrieval slower for bilinguals? Evidence from picture naming. Biling Lang Cogn 2018; 21: 479–488.

30. Bialystok E, Craik F, Luk G. Cognitive control and lexical access in younger and older bilinguals. In: Where Language Meets Thought. Routledge, 2024, pp. 84–117.

31. Taler V, Johns B. Using big data to understand bilingual performance in semantic fluency: Findings from the Canadian Longitudinal Study on Aging. PLoS One 2022; 17: e0277660.

32. Gollan TH, Fennema-Notestine C, Montoya RI, et al. The bilingual effect on Boston Naming Test performance. J Int Neuropsychol Soc 2007; 13: 197–208.

33. Gollan TH, Garcia DL, Murillo M, et al. Sprinting in two languages: Picture naming performance of older Spanish–English bilinguals on the Multilingual Naming Test Sprint 2.0. Neuropsychology.

34. Friesen DC, Luo L, Luk G, et al. Proficiency and control in verbal fluency performance across the lifespan for monolinguals and bilinguals. Lang Cogn Neurosci 2015; 30: 238–250.

35. Mariani Elena, Monastero Roberto, Mecocci Patrizia. Mild Cognitive Impairment: A Systematic Review. J Alzheimer’s Dis 2007; 12: 23–35.

36. Gallagher M, Koh MT. Episodic memory on the path to Alzheimer’s disease. Curr Opin Neurobiol 2011; 21: 929–934.

37. Johns EK, Phillips NA, Belleville S, et al. The profile of executive functioning in amnestic mild cognitive impairment: Disproportionate deficits in inhibitory control. J Int Neuropsychol Soc 2012; 18: 541–555.

38. Reinvang I, Grambaite R, Espeseth T. Executive dysfunction in MCI: Subtype or early symptom. Int J Alzheimers Dis 2012; 2012: 936272.

39. Mueller KD, Hermann B, Mecollari J, et al. Connected speech and language in mild cognitive impairment and Alzheimer’s disease: A review of picture description tasks. J Clin Exp Neuropsychol 2018; 40: 917–939.

40. Reilly J, Peelle JE, Antonucci SM, et al. Anomia as a marker of distinct semantic memory impairments in Alzheimer’s disease and semantic dementia. Neuropsychology 2011; 25: 413–426.

41. Duong A, Whitehead V, Hanratty K, et al. The nature of lexico-semantic processing deficits in mild cognitive impairment. Neuropsychologia 2006; 44: 1928–1935.

42. Garrard P, Lambon Ralph MA, Patterson K, et al. Semantic feature knowledge and picture naming in dementia of Alzheimer’s type: a new approach. Brain Lang 2005; 93: 79–94.

43. Calabria M, Cattaneo G, Marne P, et al. Language deterioration in bilingual Alzheimer’s disease patients: A longitudinal study. J Neurolinguistics 2017; 43: 59–74.

44. Costa A, Calabria M, Marne P, et al. On the parallel deterioration of lexico-semantic processes in the bilinguals’ two languages: Evidence from Alzheimer’s disease. Neuropsychologia 2012; 50: 740–753.

45. Neveu A, Goldrick M, Kleinman D, et al. Revisiting which language declines more in Spanish-English bilinguals with Alzheimer’s disease: Longitudinal decline patterns on the multilingual naming test. Neuropsychologia 2024; 202: 108948.

46. Gard S, Saad J, Sheppard CL, et al. Monolinguals outperform bilinguals in language but not executive function in aging and cognitive impairment. Neuropsychology.

47. Bialystok E, Craik FIM, Binns MA, et al. Effects of bilingualism on the age of onset and progression of MCI and AD: evidence from executive function tests. Neuropsychology 2014; 28: 290–304.

48. Kowoll ME, Degen C, Gladis S, et al. Neuropsychological Profiles and Verbal Abilities in Lifelong Bilinguals with Mild Cognitive Impairment and Alzheimer’s Disease. J Alzheimer’s Dis 2015; 45: 1257–1268.

49. Costumero V, Marin-Marin L, Calabria M, et al. A cross-sectional and longitudinal study on the protective effect of bilingualism against dementia using brain atrophy and cognitive measures. Alzheimer’s Res Ther; 12. Epub ahead of print 2020. DOI: 10.1186/s13195-020-0581-1.

50. De Bruin A, Della Sala S, Bak TH. The effects of language use on lexical processing in bilinguals. Lang Cogn Neurosci 2016; 31: 967–974.

51. Liampas I, Folia V, Morfakidou R, et al. Language Differences Among Individuals with Normal Cognition, Amnestic and Non-Amnestic MCI, and Alzheimer’s Disease. Arch Clin Neuropsychol 2023; 38: 525–536.

52. McDonnell M, Dill L, Panos S, et al. Verbal fluency as a screening tool for mild cognitive impairment. Int Psychogeriatrics 2020; 32: 1055–1062.

53. Whitford V, Titone D. The effects of word frequency and word predictability during first-and second-language paragraph reading in bilingual older and younger adults. Psychol Aging 2017; 32: 158.

54. Gale TM, Irvine K, Laws KR, et al. The naming profile in Alzheimer patients parallels that of elderly controls. J Clin Exp Neuropsychol 2009; 31: 565–574.

55. Silveri MC, Cappa A, Mariotti P, et al. Naming in patients with Alzheimer’s disease: Influence of age of acquisition and categorical effects. J Clin Exp Neuropsychol 2002; 24: 755–764.

56. Bailey LM, Lockary K, Higby E. Cross-linguistic influence in the bilingual lexicon: Evidence for ubiquitous facilitation and context-dependent interference effects on lexical processing. Biling Lang Cogn 2024; 27: 495–514.

57. Costa A, Caramazza A, Sebastian-Galles N. The cognate facilitation effect: implications for models of lexical access. J Exp Psychol Learn Mem Cogn 2000; 26: 1283.

58. Martin CD, Nozari N. Language control in bilingual production: Insights from error rate and error type in sentence production. Biling Lang Cogn 2021; 24: 374–388.

59. Li C, Gollan TH. Cognates facilitate switches and then confusion: Contrasting effects of cascade versus feedback on language selection. J Exp Psychol Learn Mem Cogn 2018; 44: 974.

60. Cuetos F, Gonzalez-Nosti M, Martínez C. The picture-naming task in the analysis of cognitive deterioration in Alzheimer’s disease. Aphasiology 2005; 19: 545–557.

61. Catricalà E, Della Rosa PA, Plebani V, et al. Semantic feature degradation and naming performance. Evidence from neurodegenerative disorders. Brain Lang 2015; 147: 58–65.

62. Gallant M, Lavoie M, Hudon C, et al. Analysis of Naming Errors in Healthy Aging, Mild Cognitive Impairment, and Alzheimer’s Disease. Can J Speech-Language Pathol Audiol; 43.

63. Mooijman S, Schoonen R, Goral M, et al. Why do bilingual speakers with aphasia alternate between languages? A study into their experiences and mixing patterns. Aphasiology 2025; 1–28.

64. Albert MS, DeKosky ST, Dickson D, et al. The diagnosis of mild cognitive impairment due to Alzheimer’s disease: Recommendations from the National Institute on Aging-Alzheimer’s Association workgroups on diagnostic guidelines for Alzheimer’s disease. Alzheimer’s Dement 2011; 7: 270–279.

65. McKhann GM, Knopman DS, Chertkow H, et al. The diagnosis of dementia due to Alzheimer’s disease: recommendations from the National Institute on Aging-Alzheimer’s Association workgroups on diagnostic guidelines for Alzheimer’s disease. Alzheimers Dement 2011; 7: 263–269.

66. Calabria M, Pérez Pérez J, Martínez-Horta S, et al. Language reconfiguration in bilinguals: A study with Huntington’s disease patients. Linguist Approaches to Biling 2021; 14: 459–483.

67. Folstein MF, Folstein SE, McHugh PR. Mini-mental state: apractical method for grading the mental state of patients for clinicians. J Psychiatr Res 1975; 12: 189–198.

68. Tamayo F, Casals-Coll M, Sánchez-Benavides G, et al. Spanish normative studies in a young adult population (NEURONORMA young adults project): Guidelines for the span verbal, span visuo-spatial, Letter-Number Sequencing, Trail Making Test and Symbol Digit Modalities Test. Neurol (English Ed 2012; 27: 319–329.

69. Morris JC, Heyman A, Mohs RC, et al. The consortium to establish a registry for Alzheimer’s disease (CERAD): I. Clinical and neuropsychological assessment of Alzheimer’s disease. Neurology 1989; 39: 1159–1165.

70. Snodgrass JG, Vanderwart M. A standardized set of 260 pictures: norms for name agreement, image agreement, familiarity, and visual complexity. J Exp Psychol Hum Learn Mem 1980; 6: 174.

71. Bates E, D’Amico S, Jacobsen T, et al. Timed picture naming in seven languages. Psychon Bull Rev 2003; 10: 344–380.

72. Guasch M, Boada R, Ferré P, et al. NIM: A Web-based Swiss army knife to select stimuli for psycholinguistic studies. Behav Res Methods 2013; 45: 765–771.

73. Forster KI, Forster JC. DMDX: A Windows display program with millisecond accuracy. *Behav Res Methods, Instruments*, Comput 2003; 35: 116–124.

74. Protopapas A. CheckVocal: A program to facilitate checking the accuracy and response time of vocal responses from DMDX. Behav Res Methods 2007; 39: 859–862.

75. Bates D, Maechler M, Bolker B, et al. Package ‘lme4’. convergence 2015; 12: 2.

76. Schwartz MF. Theoretical analysis of word production deficits in adult aphasia. Philos Trans R Soc B Biol Sci 2014; 369: 20120390.

77. Forbes-McKay KE, Venneri A. Detecting subtle spontaneous language decline in early Alzheimer’s disease with a picture description task. Neurol Sci Off J Ital Neurol Soc Ital Soc Clin Neurophysiol 2005; 26: 243–254.

78. Arce Rentería M, Casalletto K, Tom S, et al. The contributions of active spanish-english bilingualism to cognitive reserve among older hispanic adults living in California. Arch Clin Neuropsychol 2019; 34: 1235.

79. Trudeau-Meisner M, Johns BT, Taler V. Contextual diversity and picture naming: The role of aging and bilingualism. Biling Lang Cogn 2025; 1–17.

80. Svennevig J, Hansen P, Simonsen HG, et al. Code-switching in multilinguals with dementia: patterns across speech contexts. Clin Linguist Phon 2019; 33: 1009–1030.

81. Calabria M, Hernández M, Cattaneo G, et al. Active bilingualism delays the onset of mild cognitive impairment. Neuropsychologia 2020; 146: 107528.

82. Cargnelutti E, Tomasino B, Fabbro F. Effects of linguistic distance on second language brain activations in bilinguals: An exploratory coordinate-based meta-analysis. Front Hum Neurosci 2022; 15: 744489.

83. Khachatryan E, Vanhoof G, Beyens H, et al. Language processing in bilingual aphasia: a new insight into the problem. WIREs Cogn Sci 2016; 7: 180–196.

84. Kuzmina E, Goral M, Norvik M, et al. What influences language impairment in bilingual aphasia? A meta-analytic review. Front Psychol 2019; 10: 445.

