## Supplementary Material for "Naming Performance in Bilinguals with Alzheimer’s Disease and Mild Cognitive Impairment"

**Supplementary Materials: detailed statistical analyses**

1. **Accuracy analysis for CU, MCI, and AD**

Initial model for accuracy without covariates:

**Accuracy ~ Group * Log Frequency * Bilingualism type + Cognate status + (1 | ID) + (1 | ITEM)**

|  | **Accuracy** | | |
| --- | --- | --- | --- |
| *Predictors* | *Estimates* | *CI* | *p* |
| (Intercept) | 3.30 | 2.74 – 3.86 | **<0.001** |
| Group [MCI] | -1.18 | -1.65 – -0.70 | **<0.001** |
| Group [AD] | -1.30 | -1.83 – -0.77 | **<0.001** |
| Log Frequency | 0.36 | -0.00 – 0.72 | **0.052** |
| Bilingualism type [Passive] | -0.38 | -1.00 – 0.24 | 0.229 |
| Cognate status [non-cognate] | -0.20 | -0.75 – 0.35 | 0.471 |
| Group [MCI] × Log Frequency | -0.13 | -0.38 – 0.13 | 0.332 |
| Group [AD] × Log Frequency | 0.01 | -0.26 – 0.27 | 0.953 |
| Group [MCI] × Bilingualism type [Passive] | 0.79 | 0.12 – 1.47 | **0.022** |
| Group [AD] × Bilingualism type [Passive] | -0.18 | -0.93 – 0.57 | 0.635 |
| Log Frequency × Bilingualism type [Passive] | -0.25 | -0.61 – 0.12 | 0.190 |
| Group [MCI] × Freq Log × Bilingualism type [Passive] | 0.23 | -0.12 – 0.58 | 0.196 |
| Group [AD] × Freq Log × Bilingualism type [Passive] | 0.34 | -0.03 – 0.71 | 0.070 |
| AIC/BIC | 7906.5/8016.0 | | |
| Marginal R^2^ / Conditional R^2^  ^A^ | 0.066 / 0.466 | | |

2. **Accuracy: categorical frequency effect**

Extended model for accuracy with covariates and frequency as categorial variables (high vs. low)

**Accuracy ~ Group * Frequency * Bilingualism type + Cognate + MMSE + Education + Age + (1 | ID) + (1 | ITEM)**

|  | **Accuracy** | | |
| --- | --- | --- | --- |
| *Predictors* | *Estimates* | *CI* | *p* |
| (Intercept) | 3.19 | 2.56 – 3.83 | **<0.001** |
| Group [MCI] | -0.96 | -1.50 – -0.43 | **<0.001** |
| Group [AD] | -0.57 | -1.17 – 0.04 | 0.065 |
| Frequency [Low] | -0.42 | -1.13 – 0.29 | 0.249 |
| Bilingualism type [Passive] | -0.23 | -0.93 – 0.47 | 0.518 |
| Cognate status [non-cognate] | -0.12 | -0.66 – 0.42 | 0.666 |
| MMSE | 0.21 | 0.04 – 0.38 | **0.015** |
| Age | -0.33 | -0.46 – -0.21 | **<0.001** |
| Education | 0.20 | 0.07 – 0.33 | **0.003** |
| Group [MCI] × Frequency [Low] | -0.07 | -0.57 – 0.44 | 0.793 |
| Group [AD] × Frequency [Low] | -0.47 | -0.99 – 0.06 | 0.083 |
| Group [MCI] × Bilingualism type [Passive] | 0.78 | 0.05 – 1.52 | **0.037** |
| Group [AD] × Bilingualism type [Passive] | -0.08 | -0.89 – 0.74 | 0.853 |
| Frequency [Low] × Bilingualism type [Passive] | -0.15 | -0.84 – 0.55 | 0.678 |
| Group [MCI] × Frequency [Low] × Bilingualism type[Passive] | -0.14 | -0.83 – 0.54 | 0.679 |
| Group [AD] × Frequency [Low] × Bilingualism type[Passive] | -0.24 | -0.96 – 0.48 | 0.514 |
| AIC/BIC | 7633.3/7764.3 | | |
| Marginal R^2^ / Conditional R^2^ | 0.107 / 0.467 | | |

**Naming error rates as a function of word frequency (high vs. low), bilingualism type, and Group**

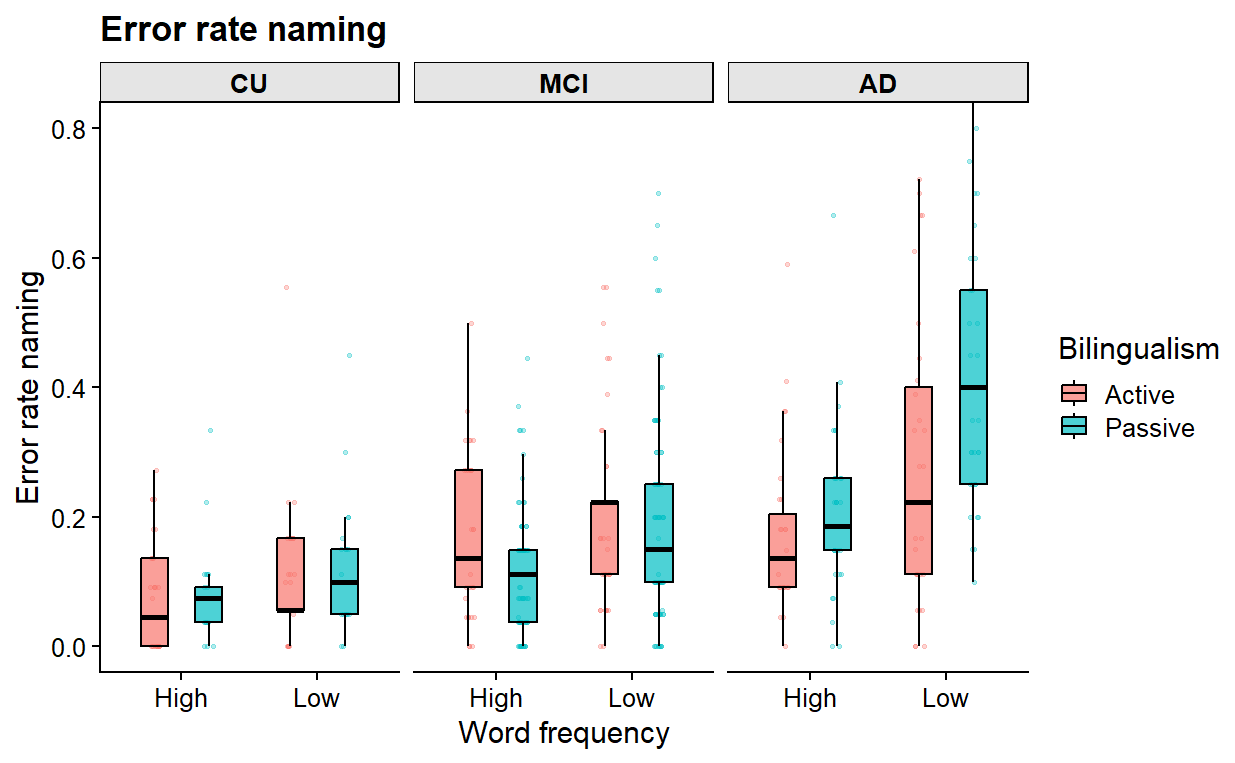

3. **Accuracy: cognate status effect**

Extended model for accuracy including only non-cognates

**Accuracy ~ Group * Log Frequency * Bilingualism type + MMSE + Education + Age + (1 | ID) + (1 | ITEM)**

|  | **Accuracy** | | |
| --- | --- | --- | --- |
| *Predictors* | *Estimates* | *CI* | *p* |
| (Intercept) | 3.39 | 2.77 – 4.00 | **<0.001** |
| Group [MCI] | -1.17 | -1.75 – -0.60 | **<0.001** |
| Group [AD] | -1.20 | -1.83 – -0.57 | **<0.001** |
| Log Frequency | 0.37 | -0.08 – 0.82 | 0.108 |
| Bilingualism type [Passive] | -0.62 | -1.33 – 0.09 | 0.088 |
| Age | -0.42 | -0.56 – -0.27 | **<0.001** |
| Education | 0.17 | 0.02 – 0.33 | **0.024** |
| MMSE | 0.26 | 0.07 – 0.44 | **0.006** |
| Group [MCI] × Log Frequency | -0.26 | -0.61 – 0.08 | 0.135 |
| Group [AD] × Log Frequency | -0.07 | -0.42 – 0.28 | 0.694 |
| Group [MCI] × Bilingualism type [Passive] | 0.85 | 0.08 – 1.62 | **0.031** |
| Group [AD] × Bilingualism type [Passive] | 0.17 | -0.67 – 1.01 | 0.688 |
| Log Frequency × Bilingualism type [Passive] | -0.29 | -0.78 – 0.20 | 0.251 |
| Group [MCI] × Log Frequency × Bilingualism type [Passive] | 0.35 | -0.14 – 0.83 | 0.163 |
| Group [AD] × Log Frequency × Bilingualism type [Passive] | 0.32 | -0.19 – 0.83 | 0.224 |
| AIC/BIC | 3796.7/3909.1 | | |
| Marginal R^2^ / Conditional R^2^ | 0.130 / 0.458 | | |

Extended model for accuracy including only cognates

**Accuracy ~ Group * Log Frequency * Bilingualism type + MMSE + Education + Age + (1 | ID) + (1 | ITEM)**

|  | **Accuracy** | | |
| --- | --- | --- | --- |
| *Predictors* | *Estimates* | *CI* | *p* |
| (Intercept) | 2.67 | 2.05 – 3.29 | **<0.001** |
| Group [MCI] | -0.81 | -1.33 – -0.29 | **0.002** |
| Group [AD] | -0.53 | -1.12 – 0.06 | 0.079 |
| Log Frequency | 0.11 | -0.50 – 0.72 | 0.729 |
| Bilingualism type [Passive] | -0.06 | -0.76 – 0.65 | 0.877 |
| Age | -0.32 | -0.46 – -0.18 | **<0.001** |
| Education | 0.16 | 0.02 – 0.31 | **0.028** |
| MMSE | 0.13 | -0.05 – 0.31 | 0.153 |
| Group [MCI] × Log Frequency | 0.32 | -0.15 – 0.78 | 0.185 |
| Group [AD] × Log Frequency | 0.46 | -0.02 – 0.94 | 0.061 |
| Group [MCI] × Bilingualism type [Passive] | 0.49 | -0.22 – 1.20 | 0.175 |
| Group [AD] × Bilingualism type [Passive] | -0.48 | -1.26 – 0.31 | 0.235 |
| Log Frequency × Bilingualism type [Passive] | 0.07 | -0.51 – 0.65 | 0.803 |
| Group [MCI] × Log Frequency × Bilingualism type [Passive] | -0.20 | -0.77 – 0.38 | 0.499 |
| Group [AD] × Log Frequency × Bilingualism type [Passive] | -0.07 | -0.67 – 0.53 | 0.817 |
| AIC/BIC | 3996.7/4108.1 | | |
| Marginal R^2^ / Conditional R^2^ | 0.082 / 0.470 | | |

**Naming accuracy in the dominant language for cognates and non-cognates**

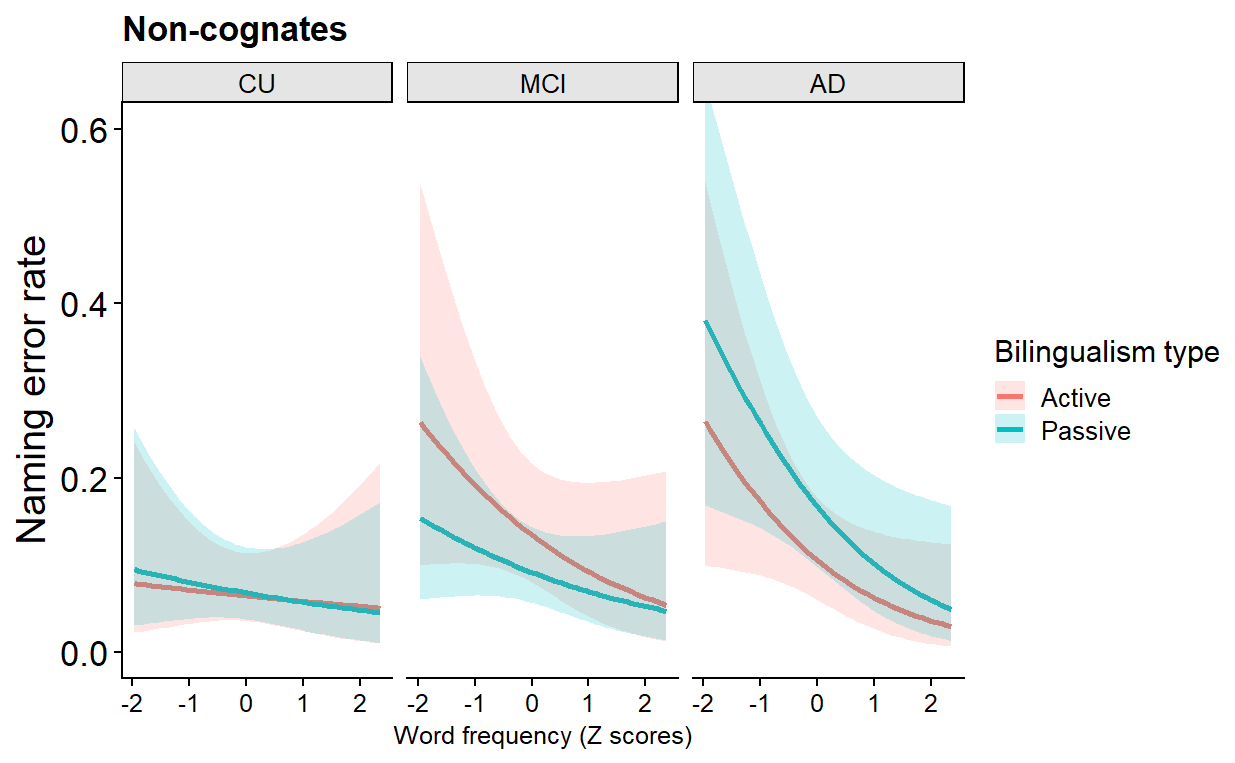

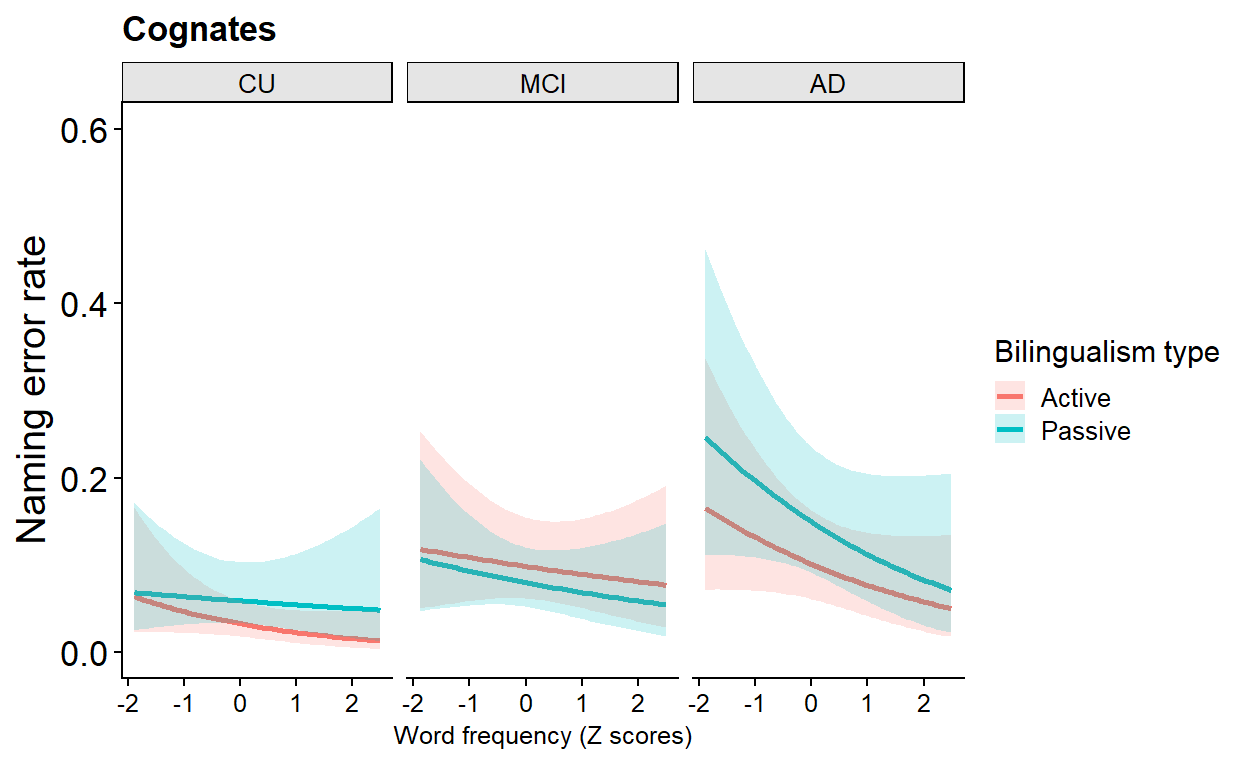

4. **Naming latencies (log-transformed RTs) analysis for CU, MCI, and AD**

Initial model for naming latencies without covariates:

**Log RT ~ Group * Log Frequency * Bilingualism type + Cognate + (1 | ID) + (1 | ITEM)**

|  | **log(RT)** | | |
| --- | --- | --- | --- |
| *Predictors* | *Estimates* | *CI* | *p* |
| (Intercept) | 7.21 | 7.15 – 7.28 | **<0.001** |
| Group [MCI] | 0.04 | -0.02 – 0.10 | 0.213 |
| Group [AD] | 0.21 | 0.13 – 0.28 | **<0.001** |
| Log Frequency | -0.02 | -0.05 – 0.01 | 0.271 |
| Bilingualism type [Passive] | 0.04 | -0.04 – 0.12 | 0.309 |
| Cognate status [non-cognate] | 0.02 | -0.04 – 0.07 | 0.523 |
| Group [MCI] × Log Frequency | -0.01 | -0.03 – 0.01 | 0.467 |
| Group [AD] × Log Frequency | 0.01 | -0.01 – 0.04 | 0.400 |
| GROUP [MCI] × Bilingualism type [Passive] | 0.01 | -0.08 – 0.10 | 0.838 |
| GROUP [AD] × Bilingualism type [Passive] | 0.08 | -0.02 – 0.19 | 0.125 |
| Log Frequency × Bilingualism type [Passive] | -0.03 | -0.07 – -0.01 | **0.033** |
| Group [MCI] × Log Frequency × Bilingualism type[Passive] | 0.00 | -0.03 – 0.04 | 0.842 |
| Group [AD] × Log Frequency x Bilingualism type [Passive] | 0.04 | 0.01 – 0.08 | **0.034** |
| AIC/BIC | 5922.9/ 6036.6 | | |
| Marginal R^2^ / Conditional R^2^ | 0.077 / 0.325 | | |

5. **Naming latencies: categorical frequency effect**

Extended model for naming latencies:

**Log RT ~ Group * Frequency * Bilingualism type + Cognate + MMSE+ Education + (1 | ID) + (1 | ITEM)**

|  | **log(RT)** | | |
| --- | --- | --- | --- |
| *Predictors* | *Estimates* | *CI* | *p* |
| (Intercept) | 7.21 | 7.14 – 7.28 | **<0.001** |
| Group [MCI] | 0.05 | -0.02 – 0.12 | 0.143 |
| Group [AD] | 0.16 | 0.07 – 0.24 | **<0.001** |
| Frequency [Low] | 0.04 | -0.03 – 0.11 | 0.256 |
| Bilingualism type [Passive] | 0.03 | -0.05 – 0.12 | 0.399 |
| MMSE | -0.03 | -0.06 – -0.01 | **0.006** |
| Age | 0.03 | 0.01 – 0.05 | **<0.001** |
| Group [MCI] × Frequency [L] | 0.01 | -0.04 – 0.06 | 0.778 |
| Group [AD] × Frequency [L] | -0.01 | -0.06 – 0.05 | 0.847 |
| Group [MCI] × Bilingualism type [Passive] | -0.03 | -0.12 – 0.07 | 0.537 |
| Group [AD] × Bilingualism type [Passive] | 0.02 | -0.09 – 0.13 | 0.732 |
| Frequency [Low] × Bilingualism type [Passive] | 0.04 | -0.02 – 0.10 | 0.226 |
| Group [MCI] × Frequency [Low] × Bilingualism type [Passive] | 0.05 | -0.01 – 0.12 | 0.119 |
| Group [AD] × Frequency [Low] × Bilingualism type [Passive] | 0.10 | 0.02 – 0.18 | **0.014** |
| AIC/BIC | 5796.1/ 5916.5 | | |
| Marginal R^2^ / Conditional R^2^ | 0.091 / 0.327 | | |

**Naming latencies (logRT) as a function of word frequency (high vs. low), bilingualism type, and Group**

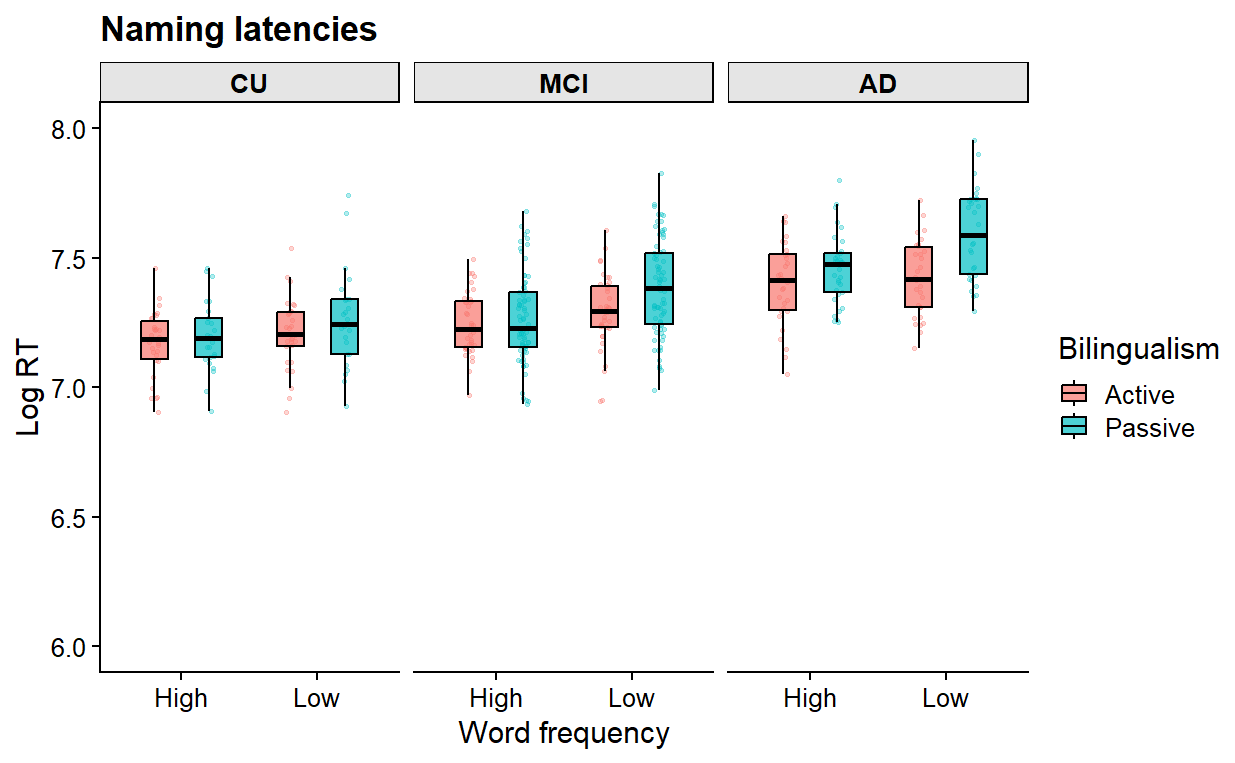

6. **Naming latencies: cognate status effect**

Extended model for naming latencies including only non-cognates

**Log RT ~ Group * Log Frequency * Bilingualism type + MMSE + Age + (1 | ID) + (1 | ITEM)**

|  | **log(RT)** | | |
| --- | --- | --- | --- |
| *Predictors* | *Estimates* | *CI* | *p* |
| (Intercept) | 7.25 | 7.18 – 7.32 | **<0.001** |
| Group [MCI] | 0.07 | -0.00 – 0.14 | 0.060 |
| Group [AD] | 0.14 | 0.05 – 0.22 | **0.001** |
| Log Frequency | -0.06 | -0.11 – -0.00 | **0.033** |
| Bilingualism type [Passive] | -0.01 | -0.10 – 0.08 | 0.896 |
| MMSE | -0.03 | -0.05 – -0.00 | **0.027** |
| Age | 0.04 | 0.02 – 0.06 | **<0.001** |
| Group [MCI] × Log Frequency | 0.01 | -0.03 – 0.05 | 0.689 |
| Group [AD] × Log Frequency | 0.02 | -0.03 – 0.06 | 0.478 |
| Group [MCI] × Bilingualism type [Passive] | -0.02 | -0.11 – 0.08 | 0.738 |
| Group [AD] × Bilingualism type [Passive] | 0.10 | -0.01 – 0.21 | 0.087 |
| Log Frequency × Bilingualism type [Passive] | -0.04 | -0.09 – 0.01 | 0.080 |
| Group [MCI] × Freq Log × Bilingualism type [Passive] | -0.00 | -0.05 – 0.05 | 0.947 |
| Group [AD] × Log Frequency × Bilingualism type [Passive] | 0.05 | -0.01 – 0.10 | 0.117 |
| AIC/BIC | 3068.9/ 3177.2 | | |
| Marginal R^2^ / Conditional R^2^ | 0.104 / 0.334 | | |

Extended model for naming latencies including only cognates

**Log RT ~ Group * Log Frequency * Bilingualism type + MMSE + Age + (1 | ID) + (1 | ITEM)**

|  | **log(RT)** | | |
| --- | --- | --- | --- |
| *Predictors* | *Estimates* | *CI* | *p* |
| (Intercept) | 7.19 | 7.12 – 7.25 | **<0.001** |
| Group [MCI] | 0.08 | 0.01 – 0.15 | **0.026** |
| Group [AD] | 0.19 | 0.11 – 0.28 | **<0.001** |
| Log Frequency | 0.02 | -0.02 – 0.06 | 0.431 |
| Bilingualism type [Passive] | 0.08 | -0.01 – 0.16 | 0.066 |
| MMSE | -0.03 | -0.06 – -0.01 | **0.010** |
| Age | 0.03 | 0.01 – 0.05 | **0.003** |
| Group [MCI] × Log Frequency | -0.02 | -0.05 – 0.01 | 0.228 |
| Group [AD] × Log Frequency | 0.00 | -0.03 – 0.03 | 0.976 |
| Group [MCI] × Bilingualism type [Passive] | -0.02 | -0.12 – 0.08 | 0.703 |
| Group [AD] × Bilingualism type [Passive] | -0.02 | -0.13 – 0.09 | 0.780 |
| Log Frequency × Bilingualism type [Passive] | -0.02 | -0.07 – 0.02 | 0.315 |
| Group [MCI] × Log Frequency ×Bilingualism type[Passive] | -0.00 | -0.05 – 0.05 | 0.991 |
| Group [AD] × Log Frequency × Bilingualism type [Passive] | 0.04 | -0.02 – 0.09 | 0.171 |
| AIC/BIC | 3045.3/ 3154.3 | | |
| Marginal R^2^ / Conditional R^2^ | 0.078 / 0.303 | | |

**Naming latencies in the dominant language for cognates and non-cognates**

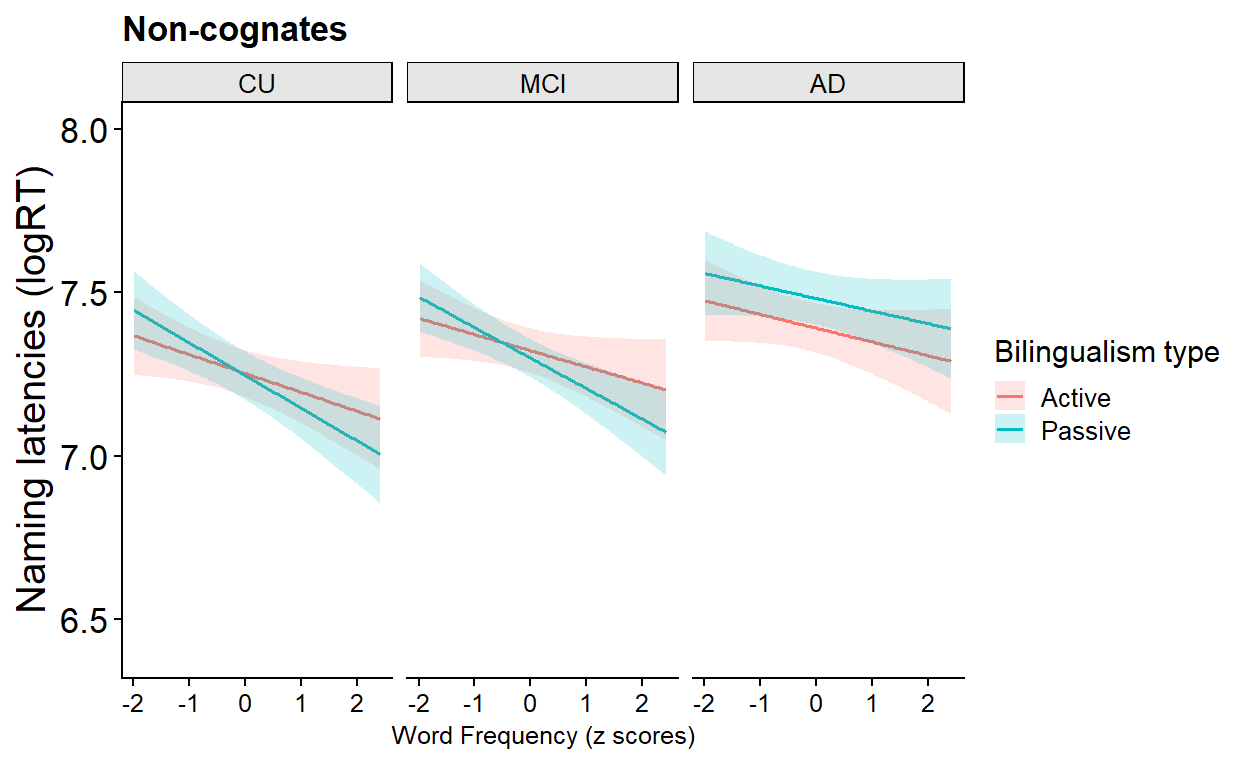

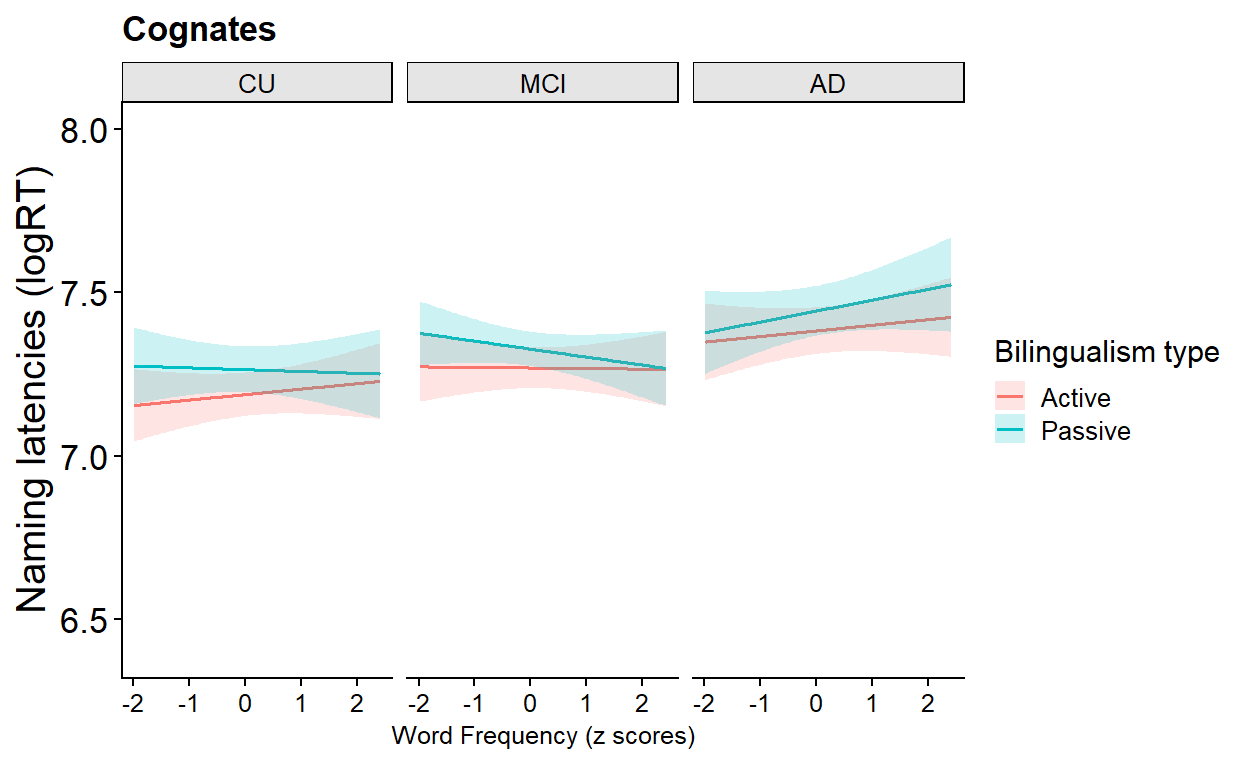

7. **Error type analysis**

Initial model for error type without covariates:

**Errors ~ Error Type * Group * Bilingualism type + (1 | ID)**

|  | **Errors** | | |
| --- | --- | --- | --- |
| *Predictors* | *Estimates* | *CI* | *p* |
| (Intercept) | 1.00 | 0.71 – 1.29 | **<0.001** |
| Error Type [Semantic] | 0.01 | -0.37 – 0.40 | 0.946 |
| Error Type [CL Intrusion] | -0.33 | -0.73 – 0.07 | 0.102 |
| Error Type [Visual] | -4.11 | -5.54 – -2.68 | **<0.001** |
| Error Type [Unrelated] | -2.60 | -3.34 – -1.86 | **<0.001** |
| Error Type [Mixed] | -1.65 | -2.17 – -1.12 | **<0.001** |
| Error Type [Other] | -3.69 | -4.87 – -2.51 | **<0.001** |
| Group [AD] | 0.51 | 0.11 – 0.90 | **0.012** |
| Bilingualism type [Passive] | -0.55 | -0.91 – -0.20 | **0.002** |
| Error Type [Semantic] × Group [AD] | -0.15 | -0.71 – 0.40 | 0.591 |
| Error Type [CL Intrusion] × Group [AD] | -1.24 | -1.89 – -0.60 | **<0.001** |
| Error Type [Visual] × Group [AD] | 0.44 | -1.33 – 2.21 | 0.625 |
| Error Type [Unrelated] × Group [AD] | -0.66 | -1.80 – 0.49 | 0.263 |
| Error Type [Mixed] × Group [AD] | -1.61 | -2.63 – -0.58 | **0.002** |
| Error Type [Other] × Group [AD] | -0.27 | -1.94 – 1.41 | 0.754 |
| Error Type [Semantic] × Bilingualism type [Passive] | 0.82 | 0.35 – 1.30 | **0.001** |
| Error Type [CL Intrusion] × Bilingualism type [Passive] | -0.74 | -1.30 – -0.18 | **0.009** |
| Error Type [Visual] × Bilingualism type [Passive] | 2.37 | 0.86 – 3.87 | **0.002** |
| Error Type [Unrelated] × Bilingualism type [Passive] | 1.24 | 0.40 – 2.09 | **0.004** |
| Error Type [Mixed] × Bilingualism type [Passive] | -0.71 | -1.51 – 0.09 | 0.083 |
| Error Type [Other] × Bilingualism type [Passive] | 0.97 | -0.41 – 2.35 | 0.167 |
| Group [AD] × Bilingualism type [Passive] | 0.57 | 0.04 – 1.09 | **0.035** |
| Error Type [Semantic] × Group [AD] × Bilingualism type [Passive] | -0.28 | -1.00 – 0.45 | 0.453 |
| Error Type [CL Intrusion] × Group [AD]) × Bilingualism type [Passive] | 0.52 | -0.38 – 1.43 | 0.254 |
| Error Type [Visual] × Group [AD]) × Bilingualism type [Passive] | -0.87 | -2.78 – 1.05 | 0.376 |
| Error Type [Unrelated] × Group [AD] × Bilingualism type [Passive] | -0.25 | -1.61 – 1.10 | 0.713 |
| Error Type [Mixed] × Group [AD] × Bilingualism type [Passive] | 0.59 | -0.88 – 2.07 | 0.430 |
| Error Type [Other] × Group [AD] × Bilingualism type [Passive] | -1.09 | -3.26 – 1.08 | 0.325 |
| AIC/BIC | 3169.9/3325.6 | | |
| Marginal R^2^ / Conditional R^2^ | 0.710 / 0.748 | | |

8. **Correlational analysis between log RT and accuracy**

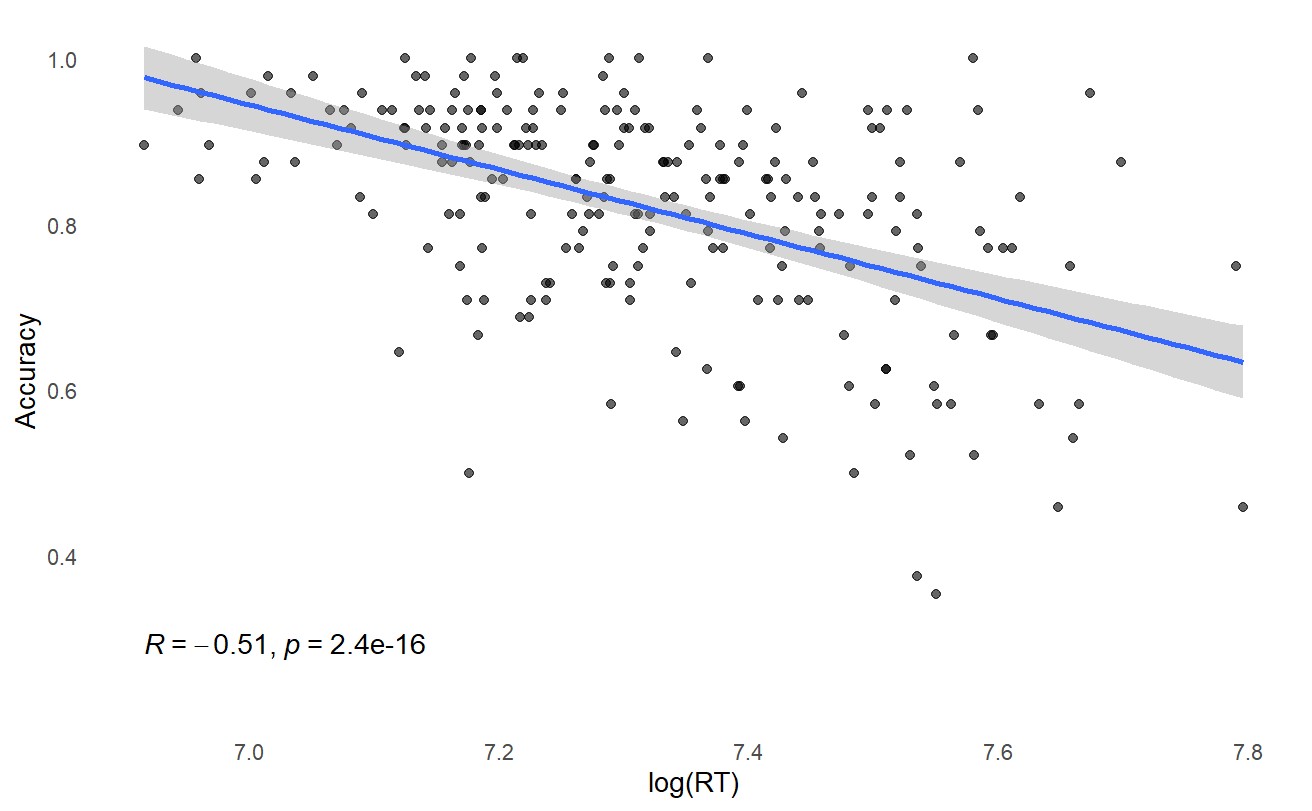

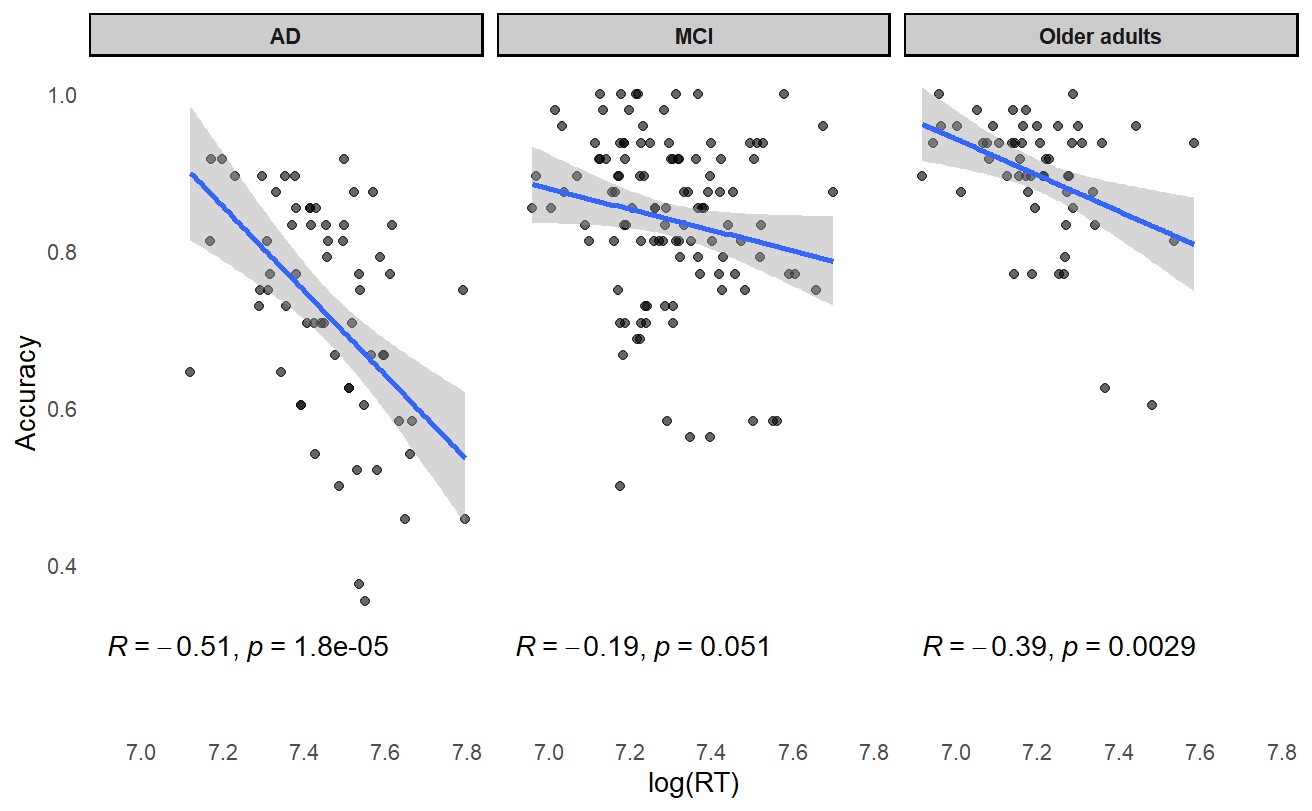
